# Predicting sensitivity and resilience to modifiable risk factors for cardiometabolic morbidity and mortality

**DOI:** 10.1101/2021.10.14.21264994

**Authors:** Hugo Pomares-Millan, Naemieh Atabaki-Pasdar, Ingegerd Johansson, Alaitz Poveda, Paul W. Franks

## Abstract

**Background:** Lifestyle exposures play a major role in the development of disease, yet people vary in their susceptibility. A critical step towards precision medicine is identifying individuals who are resilient or sensitive to the environment, and, assess whether the allocation to these predicted groups are more or less likely to develop cardiometabolic disease.

**Methods:** We have used repeated data from the VHU study (n=35440) to identify sensitive and resilient individuals using prediction intervals at the 5^th^ and 95^th^ quantile. Three exposure susceptibility groups were derived per cardiometabolic score using quantile regression forests in the training dataset; next, in the validation dataset, we assessed the different risks of the groups using Cox proportional hazard models for CVD and diabetes.

**Results:** The results of our study suggest that, after ∼10 y of follow-up, individuals with sensitivity to the environmental exposures associated with systolic and diastolic blood pressure, blood lipids, and glucose were at higher risk of developing cardiometabolic disease. Moreover, when hazards were pooled with the replication cohort, for those individuals sensitive to the exposures associated with blood pressure traits, the hazards remained significant.

**Conclusions:** Identifying individuals who are predicted to be sensitive are at higher risk of developing disease, this population may be a clinical target for prevention or early intervention and public health strategies.

## Introduction

There is growing recognition that people vary in their susceptibility to environmental risk factors for cardiometabolic diseases, suggesting that one-size-fits-all public health recommendations are unlikely to yield optimal results. Early identification of individuals who are most likely to develop diseases like type 2 diabetes (T2D) and cardiovascular disease (CVD) is desirable, as efficacious therapeutics (both lifestyle and pharmacologic) exist that can help prevent these diseases [1]. Moreover, once manifest T2D and CVD often cause life-threatening health complications can prove difficult and costly to treat [2].

Most statistical models examining susceptibility to lifestyle risk factors, from which public health recommendations are drawn, assume that a given lifestyle exposure conveys a similar effect on disease risk throughout the target population, with variability in these effects viewed solely as a consequence of measurement error and other stochastic factors [3]. However, some of this variability is likely to reflect real differences in exposure-outcome effects, such that for a lifestyle exposure like smoking, physical inactivity or sugar consumption, a gradient of risk throughout the population will exist, with some people being highly sensitive to the adverse effects of these exposures, whilst others will be resilient.

Identifying the sub-populations of those who are environmentally “sensitive” and “resilient” may have utility for the targeted prevention of cardiometabolic disease, with those considered sensitive being the target of early preventive interventions [1, 4], while alternative public health approaches may be warranted in those who appear resilient. Here we used a machine learning approach [5] to differentiate error from true variability in susceptibility to lifestyle risk factors for T2D and CVD. In so doing we sought to identify population subgroups of sensitive and resilient individuals and determine whether allocation to these subgroups affects risk of incident disease and premature mortality. The study was performed in a large, repeated measures, prospective cohort study (n >35,000) in which detailed assessments of lifestyle exposures, multiple cardiometabolic risk markers, and clinical events were obtained.

## Materials and Methods

### Study design and participants

The Västerbotten Health Survey (*Västerbottens hälsoundersökning*; VHU) [6, 7] is a prospective, population-based cohort study designed to monitor and improve health of the general population in Västerbotten county, northern Sweden. Adults residing in Västerbotten have been invited to attend their primary care centre to undertake a baseline clinical examination and complete detailed lifestyle questionnaires during the years of their 40^th^, 50^th^, and 60^th^ birthdays. We used data derived from VHU (n= 42,887) in our analyses. Owing the small subgroup of participants who were not born in Sweden (n= 7,039), the current analysis focused only on the Swedish-born contingent of VHU. Furthermore, participants in whom diabetes or cardiovascular disease had been diagnosed at baseline (n= 408) were also removed to minimize biases that can occur when people with disease diagnoses are asked to self-report their lifestyle behaviours. Participants with two health examinations between 1985 through 2016 (with ∼10 year apart between each visit) were included in the final dataset, which comprised 35,440 participants (see flowchart in Figure 1).

**Figure 1.**
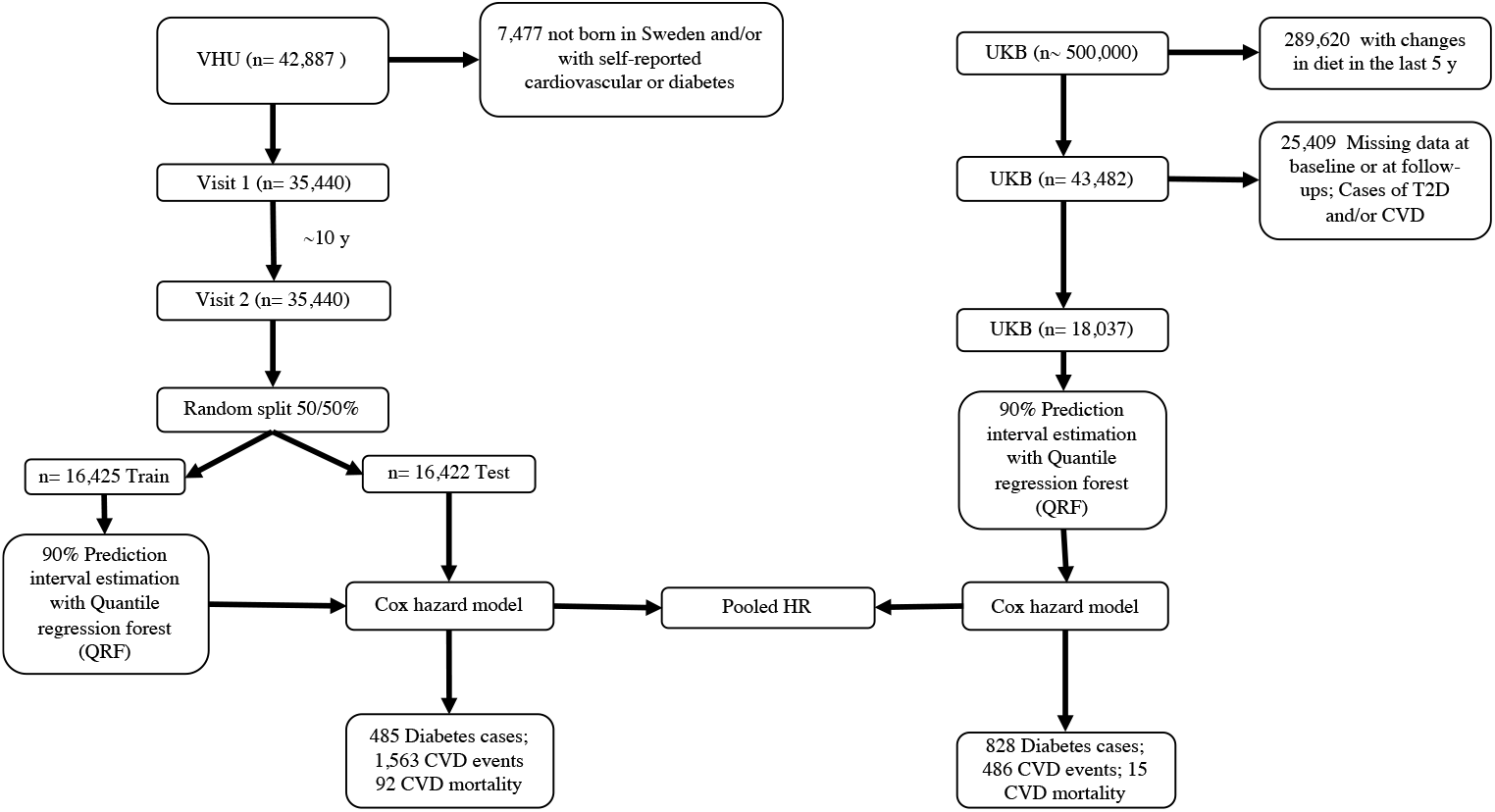
Overview of VHU and UKB studies, data processing and model training.

### Cardiometabolic risk markers

Clinical measurement protocols in VHU are reported elsewhere [6]. Briefly, height and weight were measured with calibrated tools (stadiometer and scales respectively), with participants wearing light clothing and no shoes. Body mass index (BMI) was calculated as body weight in kilograms divided by height in meters squared. Systolic and diastolic blood pressures (SBP and DBP, respectively) were measured with participants resting supine, using either manual or automated sphygmomanometers. Peripheral blood was drawn after overnight or 4-hour fasting, and a venous blood sample was drawn two hours after the administration of a 75-gram oral glucose load. Blood glucose [i.e., fasting (FG) and tow-hour glucose (2hG)], total cholesterol (TC) and triglycerides (TG) levels were then measured using a Reflotron bench-top analyser (Roche Diagnostics Scandinavia AB); High-density lipoprotein cholesterol (HDL-C) was also measured in a subgroup of participants (n= 5,348) and low-density lipoprotein cholesterol (LDL-C) obtained using the Friedewald formula (n= 5,246) [8]. In September 2009, blood lipids and blood pressure measurements changed, thereafter, blood pressure was measured twice in a sitting position and averaged; TG and TC levels were analysed using standardized chemical analysis in the hospital clinical biochemistry laboratory. Validated conversion equations were used to adjust the blood pressure and lipids measurements taken before and after September 2009 [9]: for participants on lipid lowering and/or blood pressure lowering medications, lipid levels and/or blood pressure levels were corrected by adding published constants (+0.208 mmol/l for TG, +1.347 mmol/l for TC, -0.060 mmol/l for HDL-C, +1.290 mmol/l for LDL-C, +15 mmHg for SBP and +10 mmHg for DBP) suggested in the literature [10, 11]. Cardiometabolic traits’ values outside the thresholds suggested by VHU data managers were considered outliers and removed (criteria in Supplementarytable 1).

### Lifestyle and dietary assessments

All participants were requested to complete a self-administered comprehensively validated lifestyle questionnaire during each visit; the questionnaire queries socio-economic factors, physical/mental health, quality of life, social network and support, working conditions, and alcohol/tobacco use. Physical activity was interrogated using the modified version of the International Physical Activity Questionnaire [12, 13]. A validated semi-quantitative food frequency questionnaire (FFQ) designed to capture habitual diet over the last year was used to obtain information on various dietary factors [14]. In 1996, the FFQ was reduced from 84 to 66 items by merging similar items and removing those redundant. Nutrient and energy contents were calculated based on the Swedish Food Composition Database [15] based on meal frequency and portion size. Food intake level (FIL) was calculated as total energy intake (TEI) divided by estimated basal metabolic rate, individuals with extreme TEI (below the 5th and above the 97.5th percentile of food intake level in VHU) were excluded from the analyses [16]. Observations with lifestyle values considered biologically implausible were removed (see criteria in Supplementary table 2).Written informed consent was obtained from all living participants at enrolment into VHU. The study was approved by the Region Ethical Review Board in Umeå.

### Outcome assessment

Data pertaining to medical diagnoses and mortality were retrieved through record linkage from the National Board of Health and Welfare in Sweden (www.socialstyrelsen.se/register) until December 31^st^, 2016. Using the participant’s civic registration numbers, their records were linked, and the following diagnoses codes were retrieved: ICD-9 code 250 and ICD-10 codes E11.0–E11.9 for T2D; for the composite CVD outcome: ICD-9 code 410 and ICD-10 code I21 were used for myocardial infarction (MI), and ICD-9 codes 430, 431, and 433–436 and ICD-10 codes I60, I61, I63 and I64 for stroke. The first date of a registered event was selected as the outcome for the current analyses.

### Replication in UK Biobank

UK Biobank (UKB) is a prospective cohort study since 2007 with more than 500,000 adults aged 40–70 years recruited between 2006 and 2010 in 22 centres across England, Scotland, and Wales [17]. We used the baseline assessment, which involved physical measures, sample collection, and touchscreen questionnaires that included demographics, lifestyle, environmental exposures, and medical history, as described in detail elsewhere [17]. We retrieved the same 9 traits as in VHU, with one exception: HbA1c was available in UKB, but not 2hG (the latter of which was available in VHU). The sample was restricted to individuals who reported *No* to the question: “*Have you made any major changes to your diet in the last 5 years?”* (n= 289,620). We selected those with outcome data in at least one follow-up (i.e., second or third instance) and without secondary withdrawal of consent (n= 43,482), we excluded cases of prevalent T2D, gestational diabetes or CVD, and missing data at baseline; thus, 18,037 participants were included in the analysis.

The data processing was the same as described for VHU. Dietary information was collected via a self-reported dietary questionnaire (Oxford WebQ) [18]. Outcome data, i.e., CVD incidence *(“Has a doctor ever told you that you have had any of the following conditions: angina, heart attack, stroke, high blood pressure?* at follow-up) and diabetes incidence *(“Has a doctor ever told you that you have diabetes?”* at follow-up) were self-reported. The participants were followed-up for cardiovascular mortality via linkage to information held in national death registers. Date of death was obtained from death certificates held by the National Health Service (NHS) Information Centre (England and Wales) and the NHS Central Register Scotland (Scotland). At the time of analysis, complete data from cause of death registers was available through January 31^st^, 2018; thus, they were censored at these dates or date of death if this occurred first. The field numbers for the UK Biobank variables can be viewed in the cohort’s website: http://www.ukbiobank.ac.uk/; and their correspondence with VHU variables are detailed in the supplementary material (Supplementary table 3 and 4). This research has been conducted using the data obtained via UK Biobank Access Application number 18274.

## Statistical analysis

We applied a statistical approach termed “environment-wide association study” (EWAS) for the identification of environmental risk markers, as described previously in detail [19]. Briefly, all numeric predictors were inverse normalized to correct skewness and the derived ordinal variables were treated as continuous variables in subsequent analyses. The screened predictors (∼300) were those statistically significant at the corrected *p*-value threshold after multiple testing from the EWAS. We retrieved 167 predictors for BMI, 49 and 37 for SBP and DBP, respectively, 87 for TC, 108 for TG, 50 for HDL-C and 21 with LDL-C. Forty-three predictors were associated with FG and 58 with 2hG [20]. Categorical predictors with more than two levels were converted into dummy variables, using the lowest value as reference. In VHU, reported nutrients were adjusted for TEI with the residual method [21] to obtain an unconfounded effect. We removed correlated (>80%) and zero variance predictors considered to avoid redundancy [22], variables removed are described in Supplementary table 5. For both datasets, we assumed missingness at random (MAR) [23], thus, environmental predictors with <50% missingness were imputed with *missForest* package from R software using a non-parametric approach for mixed data type, to allow a complete cases analysis suitable for the random forest algorithm; continuous predictors were verified by the mean squared error (MSE) and categorical predictors by the proportion falsely classified (PFC) (Supplementary tables 12-20)[24].

We randomly partitioned the VHU dataset into training (50%) and testing (50%) sets. The training set was used to fit quantile regression forest (QRF) models for predictors associated with the cardiometabolic traits and the testing set to predict future intervals. Multicollinearity of the variables within these models was assessed using the variance inflation factor, those >10 were removed [25]. All models were adjusted for age, age^2^, sex, FFQ version, BMI (when not as response variable), follow-up time, and fasting status (for glycaemic and lipid models). UKB models were adjusted for age, sex, BMI (when not as response variable), fasting status, and ethnicity. We utilized QRFs [5], an extension of the supervised machine learning technique random forests, which is an ensemble of simultaneous decision trees derived from bootstrapped samples [26]. Furthermore, we set prediction intervals (PIs) at 90% probability (5th and 95th quantiles) to minimize false positives [(1 - α) × 100%], while ensuring a sufficient number of events per category for the time-to-event analysis. The PIs were constructed from the conditional quantiles of the trait response predicted by QRFs. Briefly, the prediction intervals of a trait response *Y* given the environmental predictors *X* was built by *I(x)= [qα/2(Y*|*X = x), q1-α/2(Y*|*X = x)]*. Thus, the 90% prediction interval for the trait value was estimated by

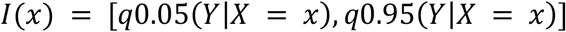

Being that for a given *x*, the trait response lies within the interval *I(x)* with high probability. For VHU, based upon the obtained PIs per trait we defined three groups of persistence when baseline visit and follow-up values fall consecutively into one of the categories: persistently ‘resilient’ (below 90% PIs), ‘as expected’ (within 90% PIs) and ‘sensitive’ (above 90% PIs). However, in UKB was not possible to consider two consecutive measures so QRFs were obtained only for the baseline visit (supplementary figures 1-9).

### Time-to-event analysis

Cox proportional hazards regression models were used to estimate hazard ratios (HRs) and corresponding 95% confidence intervals (CIs) between persistence category for each cardiometabolic trait derived from the QRF approach and the risk of diabetes and CVD-incidence and mortality. The category ‘as expected’ was used as the reference group. Statistical significance (*p-*value) was set at 5% level. Three models were constructed per trait, a basic model included age, sex, and BMI; a partially adjusted model included the basic model plus fasting status, FFQ version, TEI and educational level (education has been used as a proxy of socioeconomic status previously in this population) (22); whereas in UKB models we included ethnicity and fasting; and a third model (fully adjusted) consisted of the partially adjusted model adding smoking status, physical activity (except in UKB), and alcohol consumption. The covariates were selected *a priori* owing to their previously established associations with cardiovascular mortality in this population [27], and if a covariate was already in the environmental QRF model, it was not included. The timescale in VHU was the time-on-study and for the UKB we estimated attained age based on date of birth as the origin point (we defined the 15^th^ day of each month for all participants owing only data on month and year of births were available), date of first attendance as the entry point, and date of exit as date of death, lost to follow-up or censorship (January 31^st^, 2018). The proportional hazards assumption was tested with Schoenfeld residuals. Incidence rate (IR) ratio (cases per 100,000 person-years) and IR difference (incidence rate of ‘resilient’ or ‘sensitive’ minus incidence rate of ‘as expected’) with its 95% CIs were estimated when too few events had occurred. HRs and 95% CIs from VHU and UKB were pooled for each trait by persistence category to obtain an overall estimate under a random-effects model [28]; heterogeneity was tested with Cochran’s Q statistic [29, 30]. As sensitivity analysis, individuals in the ‘no persistence’ category were assessed in Cox proportional hazards (Supplementary table 6) and two non-modifiable covariates were added to the fully-adjusted model: *‘Parents or siblings have diabetes’* and *‘Parents or siblings had a cerebral haemorrhage/thrombosis or cardiac infarction before the age of 60’*. All statistical analyses were performed using R software version 3.6.1 [31] and packages used are listed in Supplementary table 10.

## Results

Baseline characteristics for each trait of VHU and UKB cohorts are shown in Table 1. Median follow-up [interquartile range (IQR)] for VHU was 9.7 (5.8). Fully-adjusted Cox model results per cardiometabolic trait for CVD and T2D are in table 2 and 3, respectively; basic and partially-adjusted Cox models are shown in Supplementary tables 7-9. In VHU, DBP ‘sensitive’ individuals had IRR of 2.4 (95%CI 1.5, 3.7) and the HRs were 2.25 (95%CI 1.49, 3.4; p= 1.05E-04) for CVD, conversely, the ‘resilient’ group had lower IRR of 0.4 (95%CI 0.2, 0.9) and hazards 0.39 (95%CI 0.17, 0.87; p= 2.1E-02) when compared with the reference group. For LDL-C, the IRR was 1.5 (95%CI 1.1, 3.8) in the ‘sensitive’ group and exhibited higher HRs for CVD when compared with the reference, 2.17 (95%CI 1.17, 4; p= 0.01); for TC, those ‘sensitive’ had IRR 2.5 (95%CI 1.8, 3.5) and HR 2.2 (95%CI 1.6, 3.1; p= 2.00E-06) for CVD and ∼2 times the rate (95%CI 1, 3.5) of T2D, which was also observed in UKB. In UKB cohort, the FG ‘sensitive’ group had higher rates IRR 3.5 (95%CI 2, 5.6) and risk of developing T2D than the reference, HR: 4.4 (95%CI 2.58, 7.51; p= 5.8E-08); the risk of T2D in the ‘resilient’ group was lower, yet not statistically significant.

**Table 1.** Baseline characteristics of VHU and UKB cohorts.

|  | VHU | UKB |
| --- | --- | --- |
| **n** | 35440 | 18037 |
| **Male (%)** | 15599 (46.8) | 7881 (43.7) |
| **Age (y)** | 42.96 (7.02) | 55.32 (7.58) |
| **BMI (kg/m<sup>2</sup>)** | 25.10 (3.71) | 26.02 (3.98) |
| **TC (mmol/L)** | 5.47 (1.14) | 5.88 (1.00) |
| **HDL-C (mmol/L)** | 1.32 (0.57) | 1.53 (0.37) |
| **LDL-C (mmol/L)** | 3.92 (1.16) | 3.69 (0.76) |
| **TG (mmol/L)** | 1.32 (0.76) | 1.60 (0.86) |
| **FG (mmol/L)** | 5.31 (0.63) | 4.91 (0.58) |
| **2hG (mmol/L)** | 6.39 (1.30) | - |
| **HbA1c (mmol/mol)** | - | 34.77 (3.67) |
| **SBP (mmHg)** | 123.27 (15.77) | 133.07 (15.00) |
| **DBP (mmHg)** | 77.25 (10.86) | 78.65 (8.20) |

All values are mean (SD) otherwise stated. VHU: Västerbotten Health Survey; UKB: UK biobank; BMI: Body Mass index; HDL-C: High-density lipoprotein cholesterol; LDL-C: low-density lipoprotein cholesterol; HbA1c: Glycated haemoglobin; 2hG: 2h glucose tolerance; FG: Fasting glucose; TG: Triglycerides; TC: Total cholesterol.

**Table 2.**
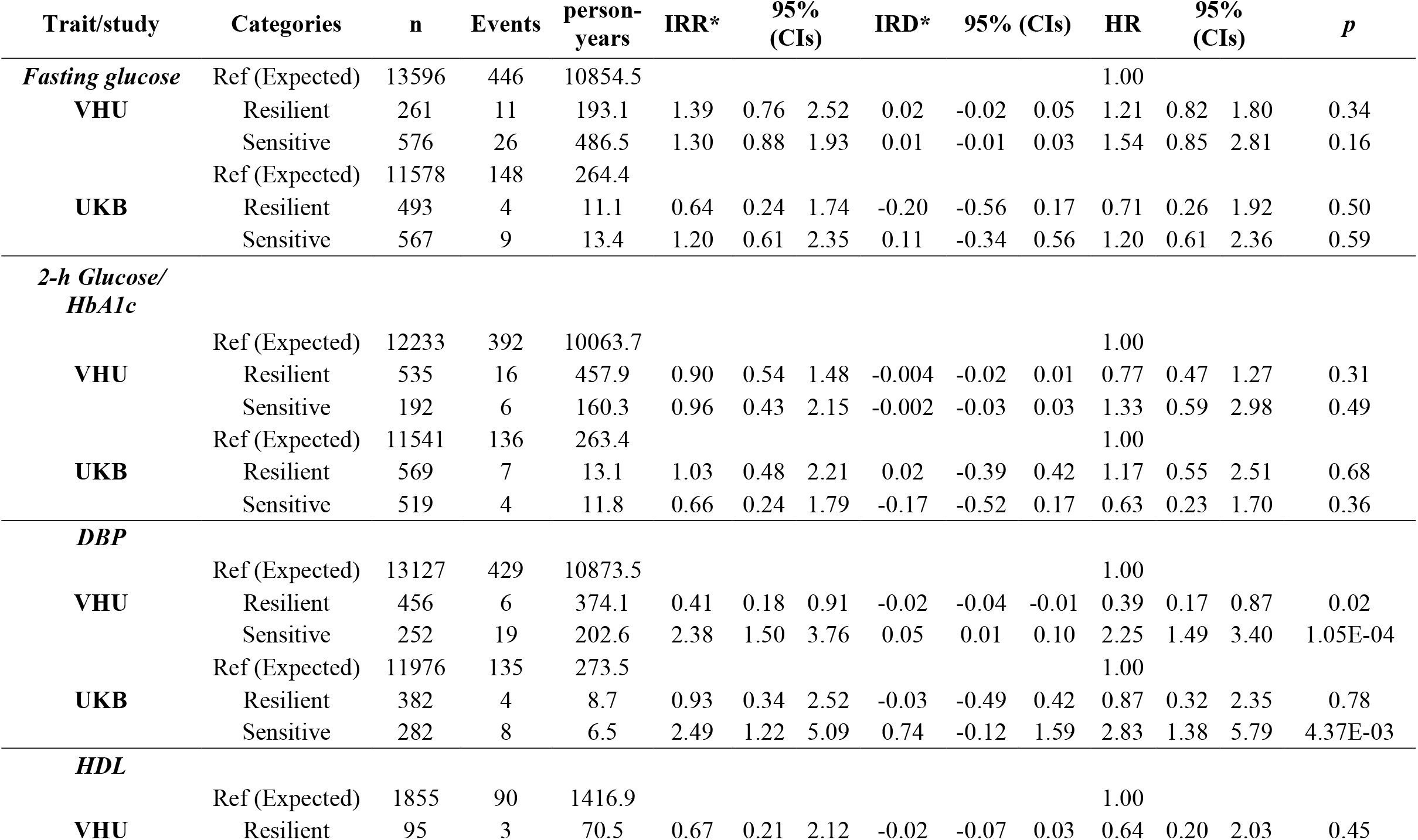
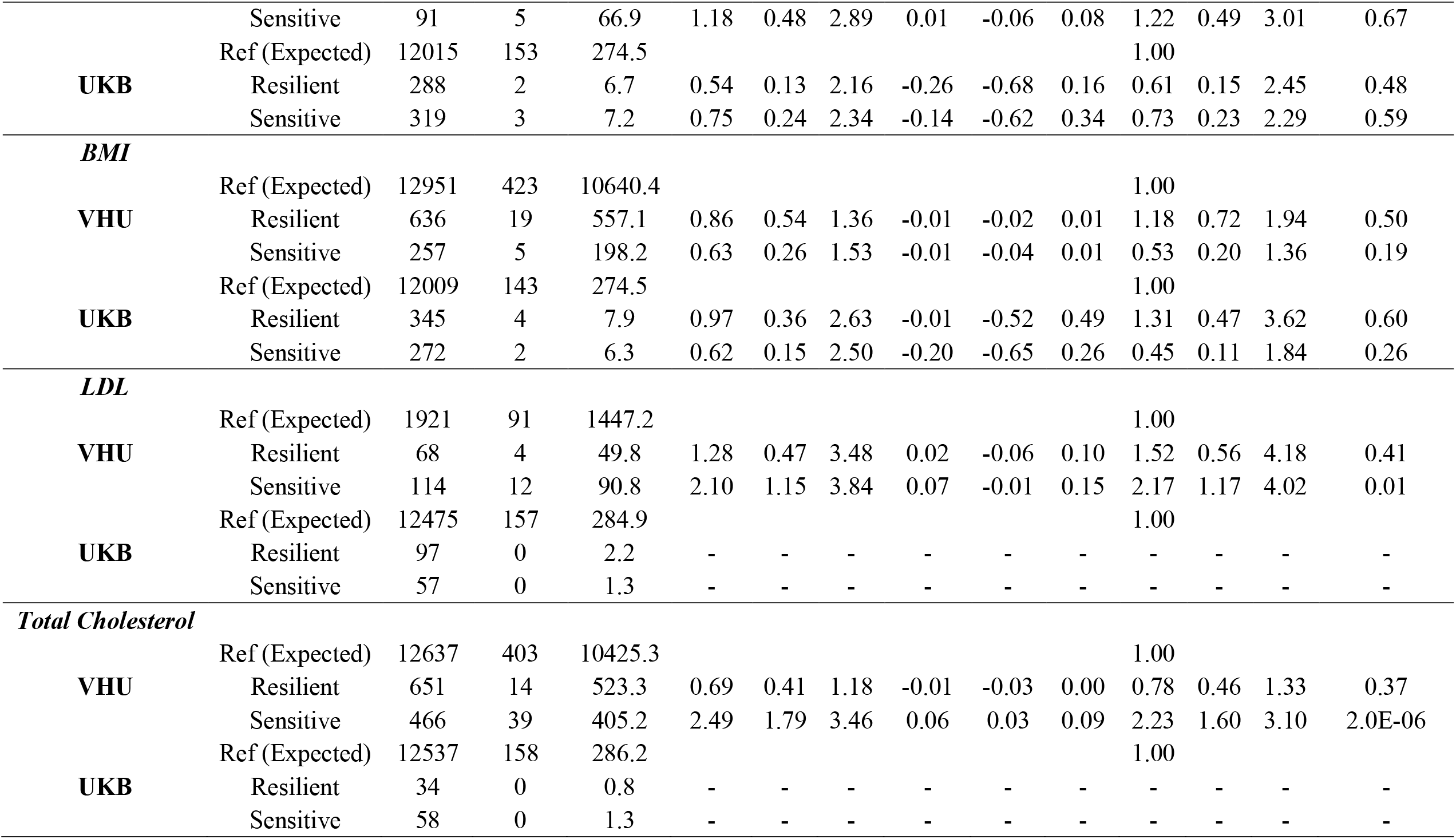
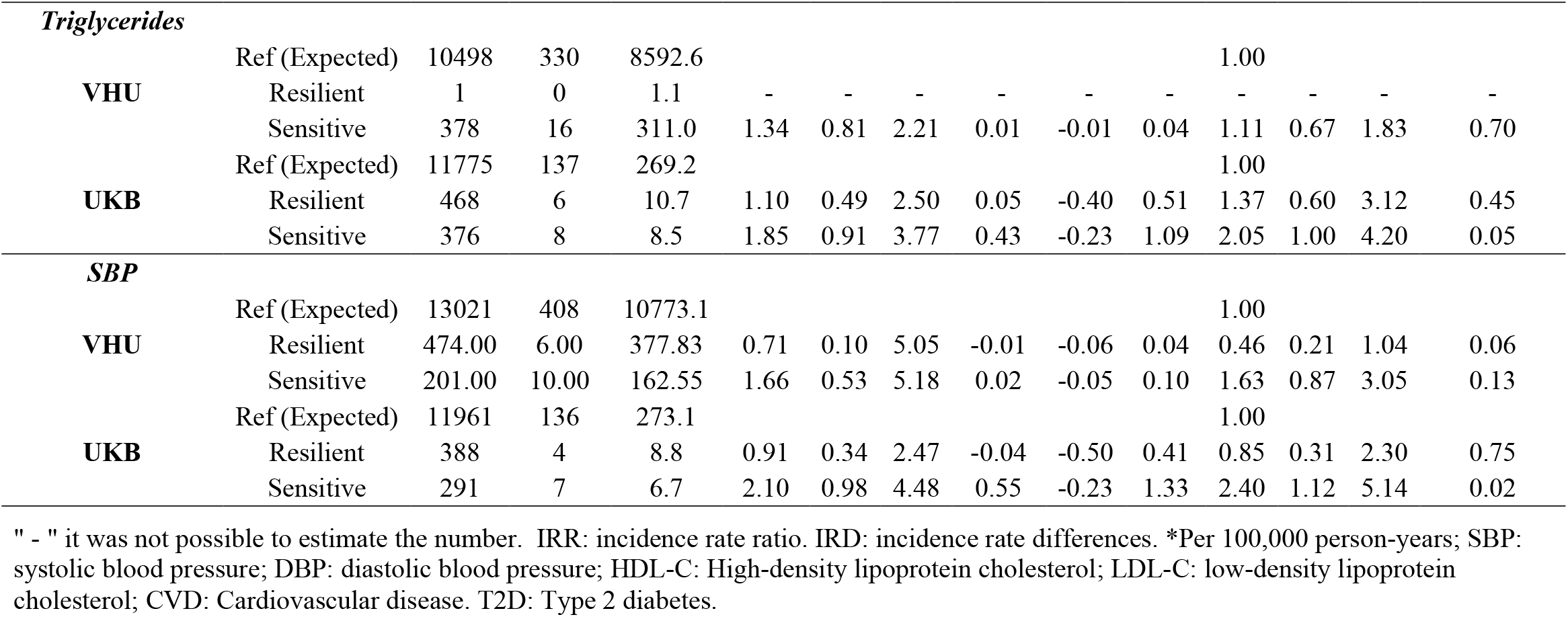
Fully-adjusted Hazard ratios and 95%CI of prediction interval categories and CVD.

**Table 3.**
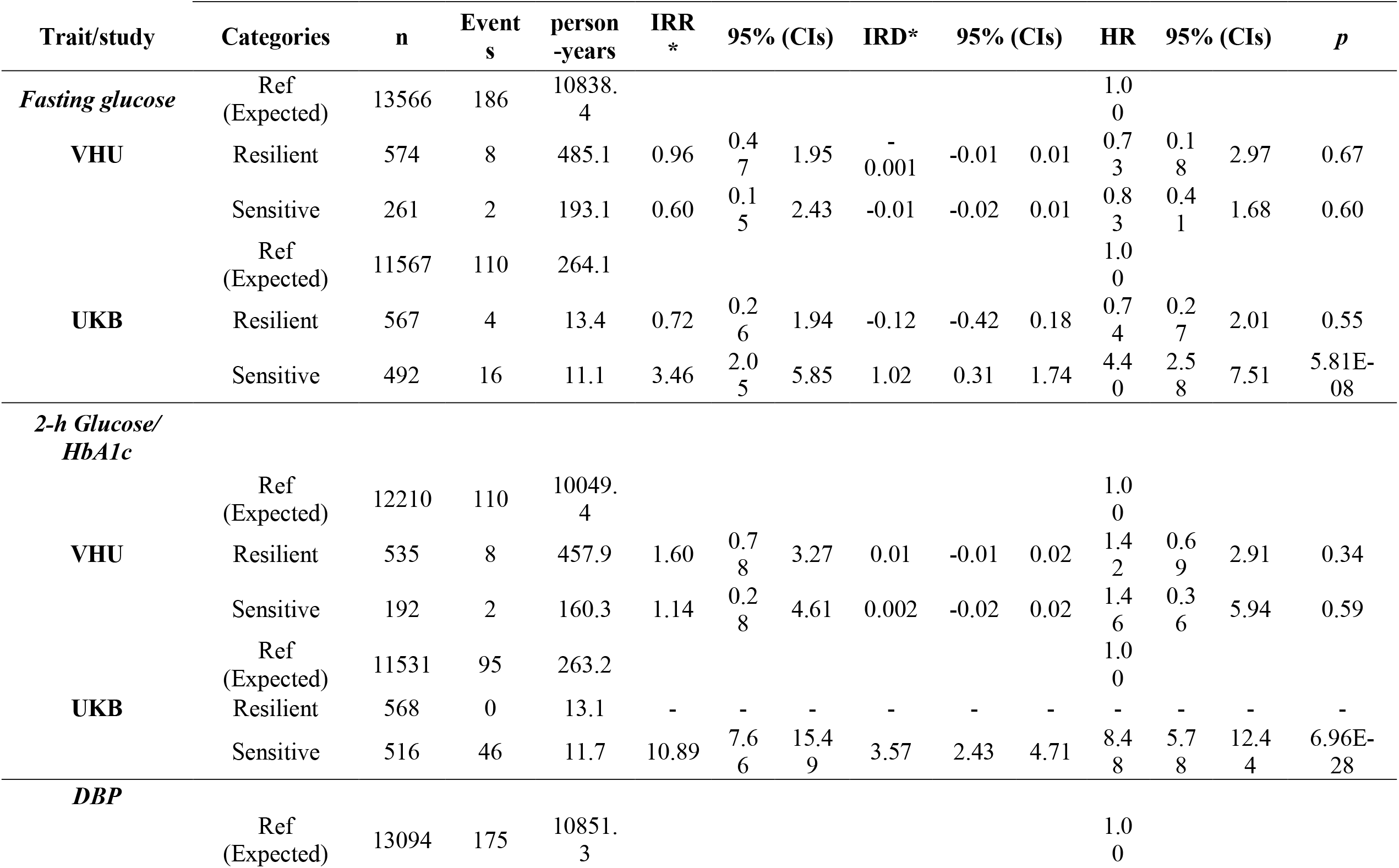
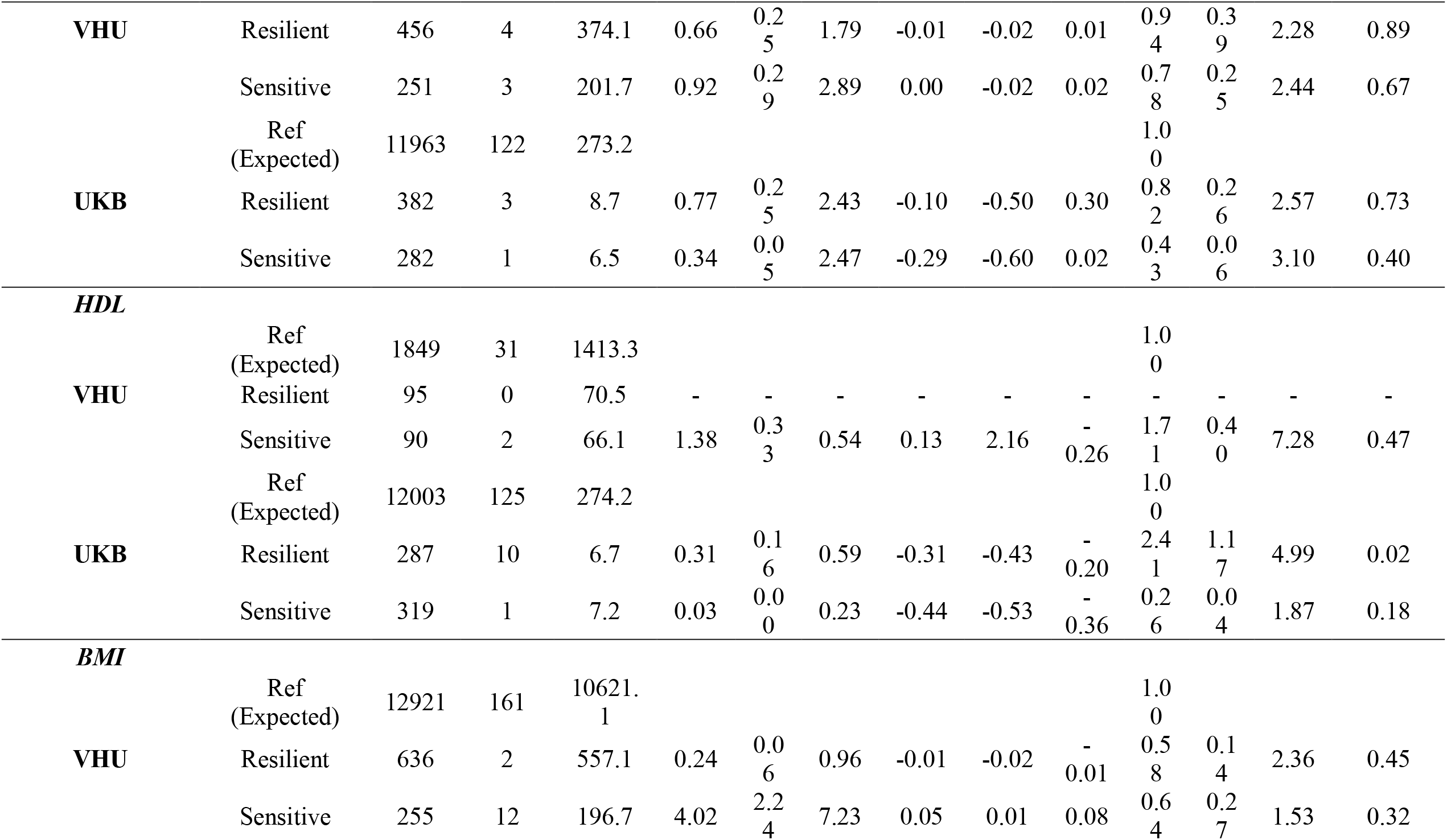
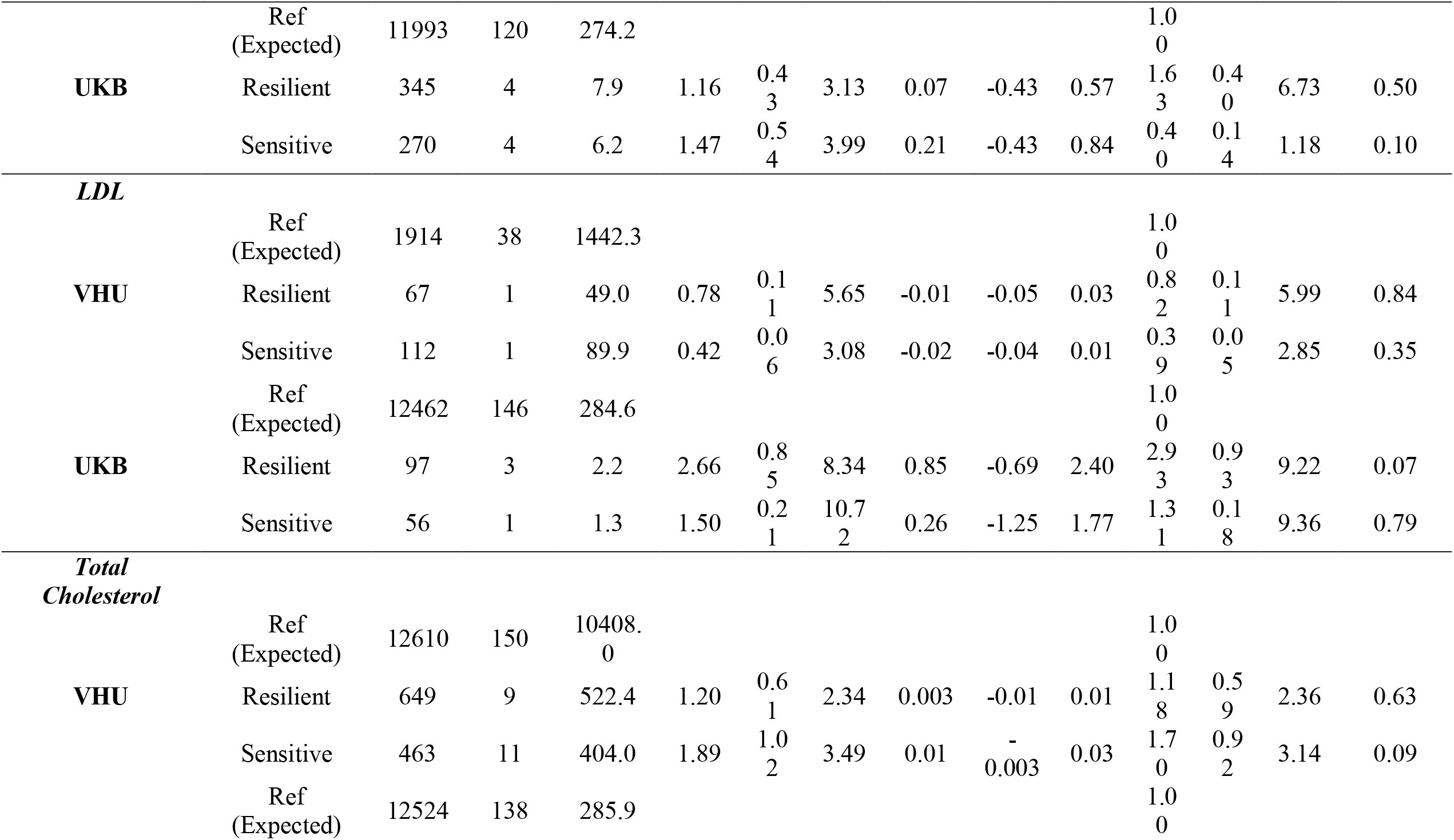
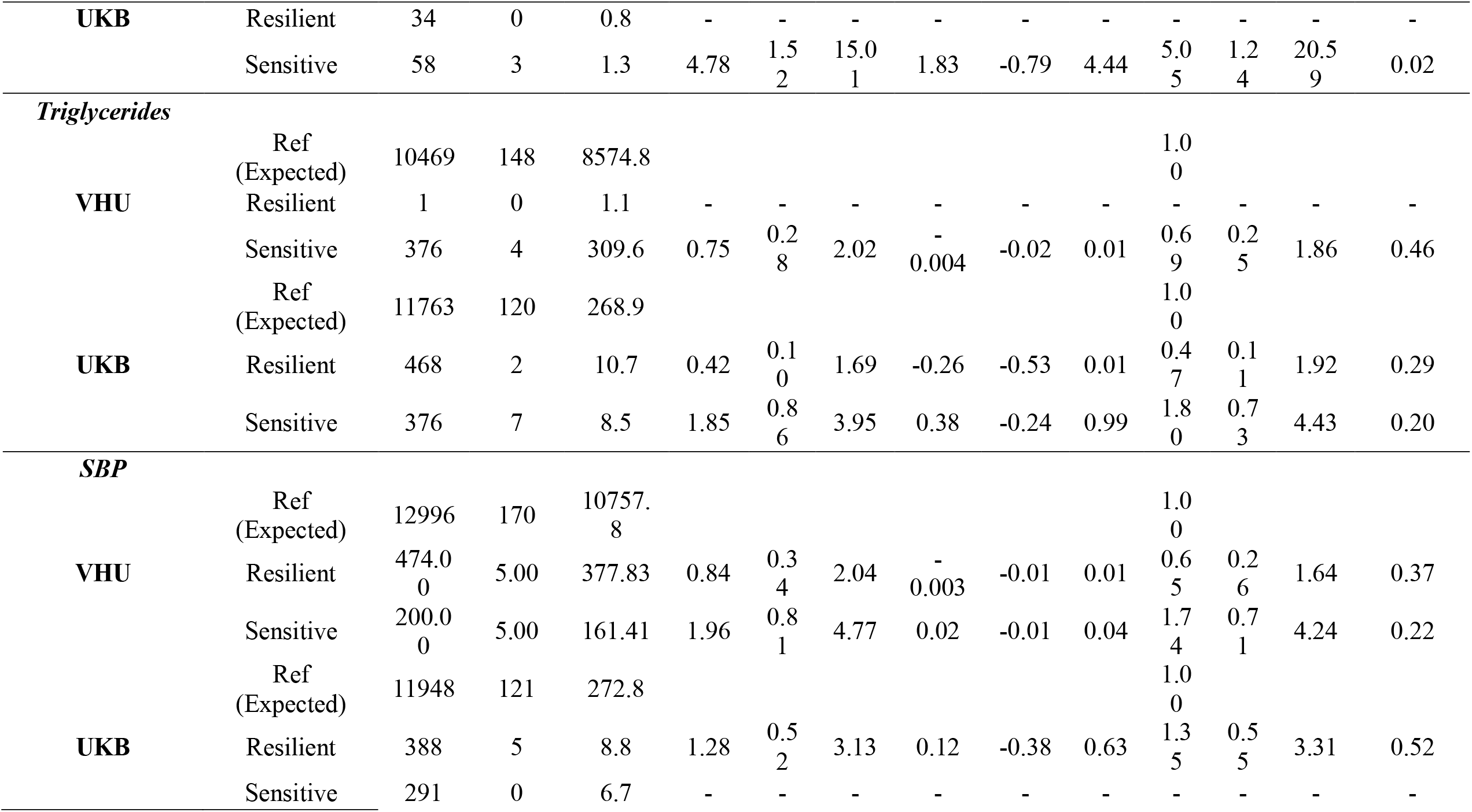
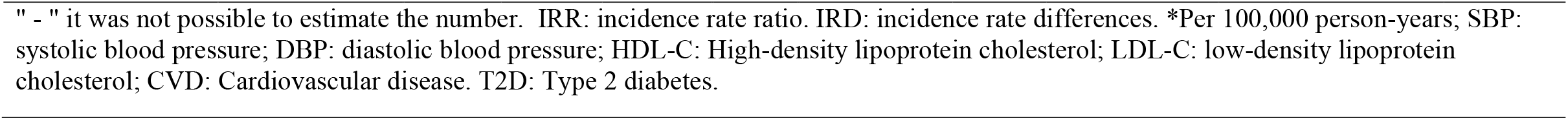
Fully-adjusted Hazard ratios and 95%CI of prediction interval categories and T2D Fully-adjusted Hazard ratios and 95%CI of prediction interval categories and T2D.

For the SBP ‘sensitive’ group, the HRs for CVD were 2.4 (95%CI 1.12, 5.14; p= 0.02) compared with the reference group, moreover, when HRs from both cohorts were pooled the overall SBP ‘sensitive’ HR for CVD was 1.9 (95%CI 1.1; 3.0), Cochran’s Q test= 8.42, *PQ*=0.0037 (Fig. 2A); Similarly, in DBP ‘sensitive’ individuals had IRR 2.5 (95%CI 1.2, 5) and HR for CVD was 2.8 (95%CI 1.38, 5.79; p= 0. 4.37E-03) and, when aggregated with VHU, the pooled DBP ‘sensitive’ HR for CVD was 2.3 (95%CI 1.5; 3.4), Cochran’s Q test= 13.3, *PQ*= 0.0003 (Fig. 2B).

**Figure 2.**
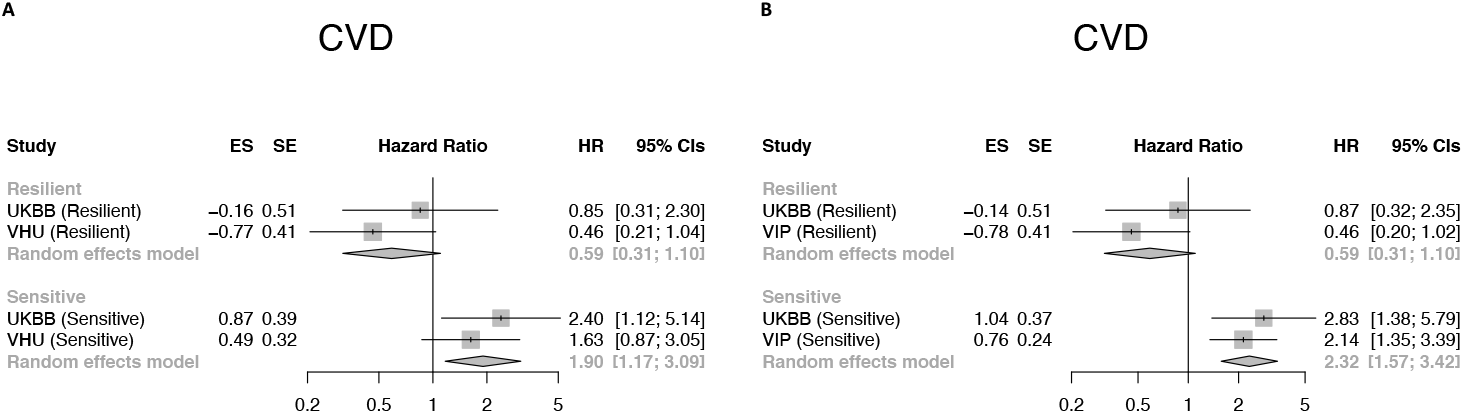
Forest plots of VHU and UKB studies by persistence category and CVD event. Panel A: Systolic blood pressure [SBP (mm/Hg)]; Panel B: Diastolic blood pressure [DBP (mm/Hg)]. Random-effects meta-analysis of the association between trait-persistence category and CVD. The square and diamond shapes represent summary estimates while the horizontal bars represent the 95% confidence intervals. HR: Hazard ratio.ES: effect estimate. SE: standard error.

In UKB, the HDL-C ‘resilient’ group were at higher hazards for T2D, 2.4 (95%CI 1.17, 4.99; p= 0.02) and those ‘sensitive’ had IRR 0.03 (95%CI 0.001, 0.23) and IRD -0.44 (95%CI -0.5, -0.4) when compared with the reference. For BMI, no fully-adjusted Cox model was significant yet the IRR of T2D in those ‘resilient’ was 0.2 (95%CI 0.06, 0.96) and, on the contrary, 4 times more (95%CI 2.2, 7.2) the rate for the ‘sensitive’ population. In UKB, for TG the hazards for CVD in the ‘sensitive’ group were twice (HR: 2; 95%CI 1.0, 4.2; p= 0.04), this association was also observed for T2D outcome, but the statistical significance disappeared when the model was maximally adjusted. For CVD-mortality outcome there were no sufficient number of events to assess and compare among groups, thus, IRR and IRD were reported (see Supplementary table 11).

## Discussion

Public health guidelines for disease risk stratification rely on population-averaged estimates of risk factor susceptibility, often involving categorization of intermediate clinical markers such as blood pressure or serum cholesterol levels. Public health guidelines for cardiometabolic disease prevention focus on reducing levels of harmful exposures such as smoking and consumption of processed foods, as well as promoting health-enhancing behaviours such as exercise and fresh fruit and vegetable consumption. However, public health guidelines do not provide recommendations tailored to the individual’s level of risk factor susceptibility. To explore whether doing so might be of clinical value, we used machine learning methods to identify, in an agnostic manner, population subgroups that are especially sensitive or resilient to modifiable risk exposures for cardiometabolic disease. We showed that those who are especially sensitive to these risk exposures tend to develop T2D and CVD more rapidly. For example, people who are sensitive to the modifiable risk factors associated with elevated blood pressure, CVD risk was twice that seen in people who responded as anticipated (i.e., had the population average response) to these risk factors; similarly, people who were sensitive to the risk factors for elevated triglycerides, LDL, and total cholesterol concentrations were twice as likely to develop CVD or diabetes. This type of risk classification is important, as it highlights individuals with “normal” or “low” levels of intermediate clinical markers who are at relatively high risk of clinical events and who would be overlooked by conventional screening and risk-classification approaches.

Of the nine intermediate clinical markers studied here, not all provided information that led to clinically relevant stratification of risk. For example, risk factor susceptibility for obesity was not associated with CVD or diabetes in either of the UK or Swedish cohorts. Similarly, sensitivity to the risk factors for elevated 2hr glucose (only available in VHU) were not associated with risk of clinical events. The scarcity of events in subgroups for traits like, LDL and HDL cholesterol in VHU and FG, HbA1c, BMI and TC in UKB, gave relatively wide confidence intervals (Tables 2 and 3). It is likely that some of the analyses performed here are underpowered and the negative findings may, by consequence, be false positive. It is also likely that some of the risk factor exposure patterns are specific to the UK and Swedish populations studied. Furthermore, although the Swedish study is characterised by its broad array of exposure assessments, the UK cohort is less well characterised, which is a key limitation for the analysis performed here. Moreover, because total energy intake was not assessed in the UK cohort, dietary exposures cannot be fully standardized, which may lead to confounding related to body composition and other features of energy balance such as exercise. One should also consider that the exposure assessments in both cohorts were obtained through questionnaires, which are prone to reporting biases, and the observational nature of the study makes causal inference challenging. Thus, while analyses of this nature are likely to yield valid estimates of clinical risk, intervening on the modifiable exposures used to define susceptibility may not necessarily reduce risk. As with most findings generated from epidemiological studies, intervention trials are required to address this limitation.

Most current clinical guidelines for T2D and CVD discuss the importance of personalized care, but include generic healthy lifestyle recommendations [32, 33]. These recommendations do not accommodate an individual’s variability in susceptibility to environmental risk factors. There has been extensive debate about the role of precision medicine in disease prevention, which typically focuses on population subgroups with distinct risk factor and treatment response profiles, such that efficacy is maximized and costs and risk are minimized [1]. The approach described here is aligned with the objectives of precision prevention [1], by aiding the identification of people at high risk of cardiometabolic disease and helping determine which modifiable exposures to intervene on.

In conclusion, the approach to cardiometabolic risk stratification presented here may help improve the precision with which at-risk subgroups of the population are identified. In practice, the implementation of this approach would require combined assessments of modifiable risk exposures and intermediate markers of cardiometabolic risk. Calculating an individual’s level of risk using the current approach is more complicated than convention risk algorithms, because it leverages conditional probabilities; however, this could be managed through app-based assessment and decision support systems, which have proven successful elsewhere [34].

## Data Availability

The individual-level data from NSHDS is not publicly available due to privacy and confidentiality constraints of Swedish regulation, but data is available from the Department of Biobank Research, Umeå University, upon reasonable request.

## Data availability

The individual-level data from NSHDS is not publicly available due to privacy and confidentiality constraints of Swedish regulation, but data is available from the Department of Biobank Research, Umeå University, upon reasonable request.

## Acknowledgements

We thank the participants, health professionals, and data managers involved in the Västerbotten Health Survey and UK Biobank. This research was conducted using the UK Biobank resource (application ID: 18274).

## SUPPLEMENTARY INFORMATION

Table 1. VHU Criteria for exclusions on cardiometabolic traits.

Table 2. VHU Criteria for implausible values for lifestyle variables.

Table 3. VHU variable meaning.

Table 4. UKBB and VHU variable correspondence.

Table 5. Variables removed during data processing.

Table 6. ‘No persistence’ category hazard ratios (HR) and 95% CIs in VHU cohort.

Table 7. Hazard ratios (HR) and 95%CI of prediction interval categories and CVD.

Table 8. Hazard ratios (HR) and 95%CI of prediction interval categories and CVD mortality.

Table 9. Hazard ratios (HR) and 95%CI of prediction interval categories and diabetes.

Table 10. R packages used for the analyses in the current study.

Table 11. Fully-adjusted Hazard ratios and 95%CI of prediction interval categories and CVD-mortality.

Table 12 (.xlsx file). Mean squared error (MSE) and proportion falsely classified (PFC) for TC.

Table 13 (.xlsx file). Mean squared error (MSE) and proportion falsely classified (PFC) for SBP.

Table 14 (.xlsx file). Mean squared error (MSE) and proportion falsely classified (PFC) for LDL-C.

Table 15 (.xlsx file). Mean squared error (MSE) and proportion falsely classified (PFC) for BMI.

Table 16 (.xlsx file). Mean squared error (MSE) and proportion falsely classified (PFC) for HDL-C.

Table 17 (.xlsx file). Mean squared error (MSE) and proportion falsely classified (PFC) for DBP.

Table 18 (.xlsx file). Mean squared error (MSE) and proportion falsely classified (PFC) for FG.

Table 19 (.xlsx file). Mean squared error (MSE) and proportion falsely classified (PFC) for 2hG.

Table 20 (.xlsx file). Mean squared error (MSE) and proportion falsely classified (PFC) for TG.

Figure 1. Scatter plots of predicted and observed values of Fasting glucose (FG) in mmol/L.

Figure 2. Scatter plots of predicted and observed values of 2-hour glucose in mmol/L and HbA1c (UKB) mmol/mol.

Figure 3. Scatter plots of predicted and observed values of Systolic blood pressure (SBP) in mm/Hg.

Figure 4. Scatter plots of predicted and observed values of Diastolic blood pressure (DBP) in mm/Hg.

Figure 5. Scatter plots of predicted and observed values of high-density lipoprotein cholesterol (HDL-C) in mmol/L.

Figure 6. Scatter plots of predicted and observed values of low-density lipoprotein cholesterol (LDL-C) in mmol/L.

Figure 7. Scatter plots of predicted and observed values of total cholesterol (TC) in mmol/L.

Figure 8. Scatter plots of predicted and observed values of triglycerides (TG) in mmol/L.

Figure 9. Scatter plots of predicted and observed values of body mass index (BMI) in kg/m2.

