## Supplementary material for "Predicting sensitivity and resilience to modifiable risk factors for cardiometabolic morbidity and mortality"

#### SUPPLEMENTARY TABLES

**Table 1. VHU Criteria for exclusions on cardiometabolic traits**

- Height: <130 cm or >210 cm
- Weight: <35 kg
- *BMI: <15 kg/m$^2$ or >70 kg/m$^2$
- *Systolic blood pressure: <20 or >300
- *Diastolic blood pressure: <20 or >250
- *Total cholesterol: <0.5 mmol/l or >15 mmol/l
- *Triglycerides: <0.15 mmol/l or >20 mmol/l. Triglycerides values lower than 0.8 mmol/l were additionally excluded due to the sensitivity of the Reflotron benchtop analyser
- *HDL-cholesterol: <0.15 mmol/l or>7 mmol/l
- *LDL-cholesterol: Not defined. LDL cholesterol values lower than 0.5 mmol/l and higher than 13 mmol/l were excluded
- *Fasting glucose: <1 mmol/l or >25 mmol/l. Fasting glucose values lower than 2 mmol/l were additionally excluded as they were considered biologically implausible
- 2h glucose: <1 mmol/l or >35 mmol/l. 2h glucose values lower than 2 mmol/l were additionally excluded as they were considered biologically implausible

*NOTE*: * Applied to UK Biobank dataset.

**Table 2. VHU Criteria for implausible values for lifestyle variables**

- Distance to work in kilometres (one way): All answers beyond 200 km were excluded
- Grams of tobacco smoked per week: All answers equal or beyond 350 gr/week were excluded
- Arachidonic acid (ARA) intake (g/day): All answers equal or beyond 0.9 gr/day were excluded
- Eicosapentaenoic acid (EPA) intake (g/day): All answers equal or beyond 2 gr/day were excluded
- Sodium intake (mg/day): All answers equal or beyond 10000 mg/day were excluded

**Table 3. VHU variable meaning**

| VHU Variable | Meaning (units) |
|---|---|
| livskvalitet_d9 | Fitness status |
| sf_3f | Physical limitation to participate in moderately demanding activities: bending down or kneeling |
| sf_3d | Physical limitation to participate in moderately demanding activities: walking up several stairs |
| sf_3a | Physical limitation to participate in strenuous activities: running, lifting heavy objects, taking part in physically demanding sports |
| sf_1 | Self-rate of overall health |
| g5 | Everyday exercise satisfaction |
| sf_3g | Physical limitation to participate in moderately demanding activities: walking more than 2 km |
| sf_11d | Excellent health |
| halsoar | Overall state of health during the last year |
| beskbltr | Informed of having high blood pressure |
| g6 | Exercise during the last three months |
| livskvalitet_d12 | Energy status |
| sf_3e | Physical limitation to participate in moderately demanding activities: walking up one flight of stairs |
| halsojf | Overall state of health compared to other your age |
| sf_7 | Pain during the last four weeks |
| sf_3b | Physical limitation to participate in moderately demanding activities: moving a table, vacuuming, walking in the forest or gardening |
| sf_8 | How much has the pain during the last four weeks disturbed your normal work? |
| pa_index_miss | Cambridge physical activity index |
| g1_3 | Cycle to work vs passive travel to work |
| sf_11b | As healthy as anyone |
| sf_9a | For how much of the time during the last four weeks have you felt really alert and strong? |
| sf_3h | Physical limitation to participate in moderately demanding activities: walking more than a few hundred meters |
| sf_4b | Physical limitation that made you do less than you wanted during the last four weeks |

| | |
|---|---|
| kottport | Average portion size of meat/fish |
| sf_9g | For how much of the time during the last four weeks have you felt worn out? |
| sf_9e | For how much of the time during the last four weeks have you felt Full of energy? |
| protsum1_anim | Animal based protein intake (g/day) |
| potport | Average portion size of potatoes/rice/pasta |
| gramnew47 | Sausage as main dish |
| sf_3j | Physical limitation to participate in moderately demanding activities: bathing or getting dressed |
| Lig_Secsum1 | Secoisolariciresinol intake ($\mu$g/day) |
| FA204_sum1 | Arachidonic acid (ARA) intake (g/day) |
| sf_4c | Physical limitation that made you not being able to perform certain work tasks or other activities during the last four weeks |
| sf_9i | For how much of the time during the last four weeks have you felt tired? |
| NIACsum1 | Vitamin B3 intake (mg/day) |
| Lig4sumsum1 | Sum of Lariciresinol, Matairesinol, Pinoresinol, Secoisolariciresinol intake ($\mu$g/day) |
| sf_11a | Get sick more often than other people |
| sf_3i | Physical limitation to participate in moderately demanding activities: walking a hundred meters |
| fibesum1 | Fibre intake (g/day) |
| sf_4d | Physical limitation that limited your ability to perform certain work tasks or other activities during the last four weeks |
| diab_foraldrar_syskon | Parents or siblings have diabetes |
| NATRsum1 | Sodium intake (mg/day) |
| g3_a | Frequency of walking during leisure time |
| g3_b | Frequency of cycling during leisure time |
| gramnew65 | Sodas, soft drinks, juice |
| gramnew43 | Minced meat dishes |
| sjukskriven | Long-term sickness |
| g4 | Changed everyday exercise during the last year |
| Lig_Pinsum1 | Pinoresinol intake ($\mu$g/day) |

| | |
|---|---|
| Folasum1 | Folate ($\mu$g/day) |
| gramnew46 | Bacon |
| Lig_Larsum1 | Lariciresinol intake ($\mu$g/day) |
| gramnew34 | Fried potatoes, pommes frites |
| Dsum1 | Vitamin D intake ($\mu$g/day) |
| g7 | If you exercise, change in exercise habits during the last year |
| sf_3c | Physical limitation to participate in moderately demanding activities: lifting or carrying grocery bags |
| scorning | Participation in sports or physical exercise associations |
| protsum1 | Total protein intake (g/day) |
| utbild | Educational level |
| sf_11c | Worsen health in the future |
| l1_4 | Porridge w/o sandwich for breakfast vs does not breakfast at all |
| MONOsum1 | Monounsaturated fat intake (g/day) |
| MOSAsum1 | Monosaccharides intake (g/day) |
| gramnew45 | Steak, chop, e.g., |
| gramnew8 | Salad dressing with oil |
| sf_6 | Extent to what your physical and emotional health disrupted your usual social life during the last four weeks |
| l1_5 | Gruel w/o sandwich for breakfast vs does not breakfast at all |
| gramnew48 | Hamburger |
| gramnew67 | Boiled coffee |
| kolesum1 | Cholesterol intake (g/day) |
| Lig_Matsum1 | Matairesinol intake ($\mu$g/day) |
| sm_status2 | Former smokers vs non-smokers |
| sf_10 | For how much of the time during the last four weeks has your physical health or your emotional problems limited your ability to interact with others? |
| gramnew44 | Meat stew |
| gramnew62 | Low fat milk (0.5%) |
| l1_3 | Sour milk, cereals, w/o sandwich for breakfast vs does not breakfast at all |

| | |
|---|---|
| sf_5b | Emotional problems that made you do less than you wanted during the last four weeks |
| ansttyp_a | Permanent employment |
| gramnew18 | Sausage, liver pâté on bread |
| gramnew72 | Wine |
| FULLKsum1 | Whole grain intake (g/day) |
| sf_4a | Physical limitation that reduced the normal time spent at work or in other activities during the last four weeks |
| gramnew41 | Pancake, waffle, Swedish dumpling |
| protsum1_veg | Plant based protein intake (g/day) |
| Lig_Sumsum1 | Sum of all lignans intake ($\mu$g/day) |
| kolhsum1 | Carbohydrates intake (g/day) |
| Bstrsum1 | Beta-sitosterol intake (mg/day) |
| ansttyp_g | Retirement pensioner full-time |
| g3_f | Frequency of hunting or fishing during leisure time |
| B12sum1 | Vitamin B12 intake ($\mu$g/day) |
| sacksum1 | Sucrose intake (g/day) |
| gramnew31 | White cabbage, lettuce, lettuce cabbage, spinach, borecole |
| livskvalitet_d5 | Satisfaction with leisure time |
| askosum1 | Vitamin C intake (mg/day) |
| gramnew42 | Pizza |
| l5a | Eat breakfast |
| Tstrsum1 | Sum of phytosterols intake (mg/day) |
| g3_g | Frequency of picking berries or mushrooms during leisure time |
| FA160_sum1 | Palmitic acid intake (g/day) |
| selesum1 | Selenium intake ($\mu$g/day) |
| Sstrsum1 | Stigmasterol intake (mg/day) |
| socforening_e | Participation in other association |
| gramnew6 | Margarine for cooking |
| gramnew24 | Fibre cereals |

| | |
|---|---|
| gramnew30 | Tomato, cucumber |
| Lig_Equsum1 | Equol intake ($\mu$g/day) |
| alkosum1 | Alcohol intake (g/day) |
| gramnew3 | Low fat margarine on bread |
| karosum1 | beta-carotene intake (mg/day) |
| g3_c | Frequency of dancing during leisure time |
| sf_5c | Emotional problems that made you do be less thorough than usual in work or other activities during the last four weeks |
| fettsum1 | Total fat intake (g/day) |
| gramnew68 | Tea |
| MAGNsum1 | Magnesium intake (mg/day) |
| sf_2 | Self-rate of overall health compared to a year ago |
| arbtala | Possibility to speak with colleagues during breaks |
| gramnew73 | Liquor, spirits |
| sn_quantity | Number of snuff boxes per week |
| sf_5a | Emotional problems that reduced the normal time spent at work or in other activities during the last four weeks |
| B6sum1 | Vitamin B6 intake (mg/day) |
| gramnew29 | Root vegetables, carrot |
| sf_9h | For how much of the time during the last four weeks have you felt happy? |
| g2_c | Light and physically active work |
| gramnew66 | Brewed (filtered) coffee |
| sf_9c | For how much of the time during the last four weeks have you felt so depressed that nothing could cheer you up? |
| livskvalitet_d10 | Appetite status |
| sm_num_cig | Number of cigarettes smoked per day |
| gramnew26 | Berries (fresh or frozen) |
| g1_2 | Walk to work vs passive travel to work |
| gramnew33 | Boiled or baked potato |

| | |
|---|---|
| FA205_sum1 | Eicosapentaenoic acid (EPA) intake (g/day) |
| sn_time | Years using snuff |
| sf_9f | For how much of the time during the last four weeks have you felt gloomy and sad? |
| sm_duration | Years smoking |
| g2_a | Sedentary or standing work |
| FA226_sum1 | Docosahexaenoic acid (DHA) intake (g/day) |
| l1_2 | Coffee/tea and wheat buns or rusk for breakfast vs does not breakfast at all |
| gronport | Average portion size of vegetables |
| gramnew7 | Oil for cooking |
| gramnew12 | White (soft) bread, thin crisp bread |
| civil3 | Marital status: Single vs Divorced/separated |
| Lig_Syrsum1 | Syringaresinol intake ($\mu$g/day) |
| l2 | Eat lunch |
| skiftarbete | Work shifts/weekends |
| gramnew64 | Milk, sour milk (3%) |
| socforening_b | Participation in study circles |
| j10 | Suggestions that you drink less |
| ansttyp_h | Retirement pensioner part-time |
| l5b | Eat lunch |
| l1_1 | Coffee/tea and sandwich for breakfast vs does not breakfast at all |
| g9 | Amount of exercise during the last 12 months |
| gramnew69 | Light beer |
| sf_9d | For how much of the time during the last four weeks have you felt calm and serene? |
| gramnew55 | Salty fish |
| gramnew38 | Pasta |
| gramnew20 | Oatflake, whole wheat, rye or barley porridge |
| gramnew22 | Sour milk, yoghurt (3% fat) |
| g10 | Time spent in a week in moderately strenuous activities |

| | |
|---|---|
| sn_status2 | Former snuff users vs non-snuff users |
| FA170_sum1 | Heptadecanoic acid intake (g/day) |
| livskvalitet_d13 | Patience status |
| ansttyp_d | Unemployment |
| ZINCsum1 | Zinc intake (mg/day) |
| Cstnsum1 | Campestanol intake (mg/day) |
| j2 | Amount of alcohol drunk in a day |
| arbfort | Job demands to work very fast |
| gramnew32 | Mixed frozen vegetables |
| gramnew70 | Medium beer |
| gramnew16 | Soft cheese |
| Bstnsum1 | Beta-sitostanol intake (mg/day) |
| gramnew35 | Mashed potato |
| j3 | Frequency of drinking six or more glasses at the same occasion |
| retisum1 | Vitamin A intake (mg/day) |
| antal_km | Distance to work in kilometres (one way) |
| sleep_h8a | Snore during sleep |
| j1 | Frequency of alcohol consumption |
| hjartinf_foraldrar_syskon | Parents or siblings had a cerebral haemorrhage/thrombosis or cardiac infarction before the age of 60 |
| gramnew59 | Sugar, honey, marmalade, jam |
| livskvalitet_d6 | Hearing status |
| mfetsum1 | Saturated fat intake (g/day) |
| g1_4 | Irregular travel mode to work vs passive travel to work |
| livskvalitet_d15 | Sleep status |
| sleep_h7b | Risk of sleeping while watching TV |
| gramnew23 | Sour milk, yoghurt (low fat) |
| livskvalitet_d4 | Satisfaction with economy |
| j8 | Times during last year that you drink so much that you were not able to remember what you did |

| | |
|---|---|
| gramnew40 | Blota (broth + bread) |
| arbkontakt | Frequent social contacts with colleagues during work |
| livskvalitet_d3 | Satisfaction with work situation |
| FA150_sum1 | Pentadecanoic acid intake (g/day) |
| gramnew25 | Corn flakes |
| DISAsum1 | Disaccharide intake (g/day) |
| soclago | Would you say that the number of people that you meet in your everyday life is enough or would you like to meet more or fewer people? |
| arbrut | Repetitive job |
| sm_gr_tobacco | Grams of tobacco smoked per week |
| soctrost | Receive hugs to comfort and support you |
| gramnew61 | Chips, popcorn, salted nuts |
| FOSFsum1 | Phosphate intake (mg/day) |
| livskvalitet_d11 | Mood status |
| gramnew27 | Apple, pear, peach, orange, mandarin and grapefruit |
| gramnew17 | Soft whey cheese |
| Lig_Endsum1 | Enterodiol intake ($\mu$g/day) |
| gramnew9 | Cream, creme fraiche, sour cream |
| sleep_h7g | Risk of sleeping while sitting still after having lunch |
| jernsum1 | Iron intake (mg/day) |
| gramnew60 | Cookies, pastry |
| gramnew28 | Banana |
| Lig_Medsum1 | Medioresinol intake ($\mu$g/day) |
| sleep_h7f | Risk of sleeping while sitting and talking with someone |
| gramnew58 | Sweets |
| sleep_h7c | Risk of sleeping while sitting inactive in a public place |
| j6 | Times during last year that you felt you needed a drink after drinking the day before |
| gramnew19 | Meat on bread |

| | |
|---|---|
| gramnew2 | Butter on bread |
| sn_status1 | Snuff user vs non-snuff user |
| socsam | Number of social interactions during a normal week |
| l3 | Eat dinner |
| gramnew49 | White meat (poultry) |
| socstod | Support from others |
| B2sum1 | Vitamin B2 intake ($\mu$g/day) |
| g3_e | Frequency of gardening during leisure time |
| livskvalitet_d1 | Satisfaction with home and family situation |
| l1_0 | Only coffee/tea for breakfast vs does not breakfast at all |
| TRANSsum1 | Trans fat intake (g/day) |
| JODIsum1 | Iodine intake ($\mu$g/day) |
| Lig_Enlsum1 | Enterolactone intake ($\mu$g/day) |
| sambo2 | Cohabitation: Live alone vs Only children |
| graviditetsdiabetes | Had gestational diabetes |
| sochem | Number of friends that can come to your home at any time and feel at home |
| socdelta | Participation in associations or voluntary organisations |
| sleep_h7e | Risk of sleeping while lying down resting in the afternoon |
| livskvalitet_d14 | Confidence status |
| i1 | Teetotaler |
| j4 | Times during last year that you felt inability to stop drinking |
| gramnew63 | Milk, sour milk (1,5%) |
| gramnew56 | Smoked fish/meat |
| ensum1 | Total energy intake (kcal/day) |
| POLYsum1 | Polyunsaturated fat intake (g/day) |
| sleep_h7h | Risk of sleeping in a car which has stopped for a few minutes |
| Cstrsum1 | Campesterol intake (mg/day) |
| gramnew5 | Butter for cooking |

| | |
|---|---|
| socupps | Appreciation of the ones at home or others |
| livskvalitet_d16 | Do you feel important and appreciated outside your home? |
| livskvalitet_d2 | Satisfaction with accommodation |
| arbski | Skill demand from job |
| gramnew14 | Cheese 28% |
| sleep_h7a | Risk of sleeping while sitting and reading |
| FA140_sum1 | Formic acid intake (g/day) |
| sleep_h7d | Risk of sleeping as a passenger in a car for one hour without break |
| ansttyp_c | Work at home |
| FA182_sum1 | Linoleic acid intake (g/day) |
| arbvad | Control over own work assignment |
| gramnew10 | Whole grain crisp bread |
| gramnew15 | Cheese 10-17% |
| g2_b | Light but partly physically active work |
| gramnew4 | Margarine on bread |
| i3 | Receive critics about your alcohol consumption |
| sambo1 | Cohabitation: Live alone vs Only one adult (spouse, partner) |
| ansttyp_f | Self-employed |
| livskvalitet_d17 | Do you feel important and appreciated in your home? |
| arbhin | Enough time for job assignments |
| i2 | Feel the need to reduce alcohol consumption |
| i4 | Feel uneasy or guilty because of your way of drinking |
| socforening_d | Participation in choir |
| gramnew52 | Lean fish (e.g., perch, bass, cod) |
| gramnew13 | Coffee rolls/buns, rusk |
| gramnew1 | Bregott on bread |
| arbpsyk | High mental demand from job |
| j7 | Times during last year that you felt guilty because of your drinking |

| | |
|---|---|
| i5 | Drunk alcohol first thing in the morning |
| TIAMsum1 | Tiamin intake (mg/day) |
| arbhur | Control over planning and execution of the workday |
| j9 | Hurt people because of your drinking |
| arbnytt | Learn new things at job |
| gramnew37 | Rice |
| soclana | People to ask for favours from |
| soctala | Number of people with whom you can speak openly |
| sm_status5 | Former occasional smoker vs non-smoker |
| gramnew51 | Liver, kidney |
| kalcsum1 | Calcium intake (mg/day) |
| sm_status1 | Smoker vs non-smoker |
| gramnew71 | Strong beer |
| sambo4 | Cohabitation: Live alone vs Other/others |
| gramnew50 | Blood based food |
| socforening_c | Participation in theatre group |
| gramnew39 | Brown beans, pea soup |
| sockont | Number of social contacts with the same interests as you |
| sf_9b | For how much of the time during the last four weeks have you felt very nervous? |
| gramnew21 | Rosehip, sweet syrup soup |
| l5c | Eat dinner |
| ansttyp_e | Student |
| gramnew57 | Ice cream |
| arbfritid | Frequency of social contacts with colleagues during leisure time |
| civil4 | Marital status: Single vs Widow/widower |
| arbfys | High physical demand from job |
| socanfo | Person to confide in |
| sambo3 | Cohabitation: Live alone vs Adult and children |

| | |
|---|---|
| j5 | Times during last year that something was not done due to your drinking |
| KALIsum1 | Potassium intake (mg/day) |
| socnara | Close relationship with anyone |
| socofta | Frequency of engaging in clubs, associations or study circles |
| gramnew53 | Fatty fish (e.g. herring, whitefish, salmon) |
| g2_e | Physically straining work most of the time |
| gramnew54 | Shellfish (e.g. shrimps, scallops) |
| arblamna | Possibility to leave your work for a while to speak with a colleague |
| tokosum1 | Vitamin E intake (mg/day) |
| ansttyp_b | Temporary employment |
| gramnew36 | Potato salad |
| g3_d | Frequency of shovelling snow during leisure time |
| sochelp | People to ask for help apart from the ones at home |
| sm_cig_groups | Number of cigarettes smoked per day (in groups) |
| soclyck | Special person to share feelings |
| g2_d | Sometimes physically straining work |
| arbkrav | Contradictory demands in job |
| arbbesok | Last time a colleague visited you at home |
| livskvalitet_d7 | Vision status |
| sm_num_cigar | Number of cigars smoked per day |
| gramnew11 | Whole grain soft bread |
| sm_status4 | Occasional smoker vs non-smoker |
| FA183_sum1 | Linolenic acid intake (g/day) |
| arbide | Ingenuity or creativity demand from job |
| livskvalitet_d8 | Memory status |
| civil2 | Marital status: Single vs Married/partner |
| sleep_h8b | Breath-holds during sleep |
| ansttyp_i | Retirement pensioner unspecified |

**Table 4. UKBB and VHU variable correspondence**

| UKBB Field ID | UKBB Variable | VHU Variable |
|---|---|---|
| | eid | Subject_id |
| 40000 | date_of_death | date_of_death |
| 40001 | cause_icd10 | cause_icd10 |
| 31 | sex_f31_0_0 | gender |
| 74 | fasting_time_f74_0_0 | fasta_enk |
| 93 | systolic_blood_pressure_manual_reading_f93_0_0 | sbt |
| 94 | diastolic_blood_pressure_manual_reading_f94_0_0 | dbt |
| 95 | pulse_rate_during_bloodpressure_measurement_f95_0_0 | - |
| 102 | pulse_rate_automated_reading_f102_0_0 | - |
| 191 | date_lost_to_followup_f191_0_0 | - |
| 1160 | sleep_duration_f1160_1_0 | livskvalitet_d15 |
| 1170 | getting_up_in_morning_f1170_0_0 | - |
| 1180 | morningevening_person_chronotype_f1180_0_0 | - |
| 1190 | nap_during_day_f1190_0_0 | - |
| 1200 | sleeplessness_insomnia_f1200_0_0 | - |
| 1210 | snoring_f1210_0_0 | - |
| 1220 | daytime_dozing_sleeping_narcolepsy_f1220_0_0 | - |
| 1239 | current_tobacco_smoking_f1239_0_0: No vs. Yes | sm_status1<br>sn_status1 |
| 1249 | past_tobacco_smoking_f1249_0_0: No vs. Yes | sm_status2<br>sn_status2 |
| 1259 | smokingsmokers_in_household_f1259_0_0: No vs. Yes | - |
| 1269 | exposure_to_tobacco_smoke_at_home_f1269_0_0 | - |
| 1279 | exposure_to_tobacco_smoke_outside_home_f1279_0_0 | - |
| 1289 | cooked_vegetable_intake_f1289_0_0 | gramnew29<br>gronport |
| 1299 | salad_raw_vegetable_intake_f1299_0_0 | gramnew32<br>gramnew8 |
| 1309 | fresh_fruit_intake_f1309_0_0 | gramnew27 |

| | | |
|---|---|---|
| 1319 | dried_fruit_intake_f1319_0_0 | - |
| 1329 | oily_fish_intake_f1329_0_0 | gramnew53 |
| 1339 | nonoily_fish_intake_f1339_0_0 | gramnew52 |
| 1349 | processed_meat_intake_f1349_0_0 | - |
| 1359 | poultry_intake_f1359_0_0 | gramnew49 |
| 1369 | beef_intake_f1369_0_0 | - |
| 1379 | lambmutton_intake_f1379_0_0 | - |
| 1389 | pork_intake_f1389_0_0 | - |
| 1408 | cheese_intake_f1408_0_0 | gramnew17 |
| | | gramnew16 |
| | | gramnew14 |
| | | gramnew15 |
| 1418 | milk_type_used_f1418_0_0 | gramnew63 |
| 1418 | milk_type1: semi-skimmed vs. never | gramnew64 |
| 1418 | milk_type2:skimmed vs. never | gramnew62 |
| 1418 | milk_type3: full cream vs. never | gramnew62 |
| 1418 | milk_type4: soy vs. never | - |
| 1418 | milk_type5: other milk vs. never | - |
| 1428 | spread_type_f1428_0_0 | - |
| 1428 | spread_type1: Butter/spreadable butter vs. never | - |
| 1428 | spread_type2: Flora Pro-Active/Benecol vs. never | - |
| 1428 | spread_type3: "Other type of spread/margarine vs. never | - |
| 1438 | bread_intake_f1438_0_0 | gramnew40 |
| 1448 | bread_type_f1448_0_0 | gramnew10 |
| 1448 | bread_type1: Wholemeal or wholegrain vs. White | - |
| 1448 | bread_type2: Wholemeal or wholegrain vs. Brown | - |
| 1448 | bread_type3: Wholemeal or wholegrain vs. Other type of bread | - |
| 1458 | cereal_intake_f1458_0_0 | - |
| 1468 | cereal_type_f1468_0_0 | gramnew24 |
| 1468 | cereal_type1: Bran cereal (e.g., All Bran, Branflakes) vs. Biscuit cereal | - |
| 1468 | cereal_type2: Bran cereal (e.g., All Bran, Branflakes) vs. Oat cereal | - |
| 1468 | cereal_type3: Bran cereal (e.g., All Bran, Branflakes) vs. Muesli | - |

| | | |
|---|---|---|
| 1468 | cereal_type4: Bran cereal (e.g., All Bran, Branflakes) vs. Other | - |
| 1478 | salt_added_to_food_f1478_0_0 | - |
| 1488 | tea_intake_f1488_0_0 | gramnew68 |
| 1498 | coffee_intake_f1498_0_0 | - |
| 1508 | coffee_type_f1508_0_0 | gramnew66 |
| 1508 | coffee_type1: Decaffeinated coffee vs. Instant coffee | - |
| 1508 | coffee_type2: Decaffeinated coffee vs. Ground coffee | - |
| 1508 | coffee_type3: Decaffeinated coffee vs. Other type of coffee | gramnew67 |
| 1518 | hot_drink_temperature_f1518_0_0 | - |
| 1528 | water_intake_f1528_0_0 | - |
| 1538 | major_dietary_changes_in_the_last_5_years_f1538_0_0: No vs. Yes | - |
| 1548 | variation_in_diet_f1548_0_0 | - |
| 1558 | alcohol_intake_frequency_f1558_0_0 | j3 |
| | | Alcohol intake (g/day) |
| 1568 | average_weekly_red_wine_intake_f1568_0_0 | gramnew72 |
| 1578 | average_weekly_champagne_plus_white_wine_intake_f1578_0_0 | - |
| 1588 | average_weekly_beer_plus_cider_intake_f1588_0_0 | - |
| 1598 | average_weekly_spirits_intake_f1598_0_0 | gramnew73 |
| 1608 | average_weekly_fortified_wine_intake_f1608_0_0 | - |
| 1618 | alcohol_usually_taken_with_meals_f1618_0_0 | - |
| | among current drinkers, drinks usually with meals: yes + it varies vs. no | - |
| | among current drinkers, drinks usually with meals: yes vs. no | - |
| 1628 | alcohol_intake_versus_10_years_previously_f1628_0_0 | - |
| 2443 | diabetes_diagnosed_by_doctor_f2443_0_0 | - |
| 2644 | light_smokers_at_least_100_smokes_in_lifetime_f2644_0_0 | sm_status4 |
| | | sm_status5 |
| 2654 | nonbutter_spread_type_details_f2654_0_0 | - |
| 2654 | nonbutter_spread1: Olive oil based spread vs. Flora Pro-Active or Benecol | - |
| 2654 | nonbutter_spread2: Olive oil based spread vs. Soft (tub) margarine | - |
| 2654 | nonbutter_spread3: Olive oil based spread vs. Hard (block) margarine | - |
| 2654 | nonbutter_spread4: Olive oil based spread vs. Polyunsaturated/sunflower oil based spread | - |
| 2654 | nonbutter_spread5: Olive oil based spread vs. Other low or reduced fat spread | - |

| | | |
|---|---|---|
| 2654 | nonbutter_spread6: Olive oil based spread vs. Other type of spread/margarine | - |
| 2664 | reason_for_reducing_amount_of_alcohol_drunk_f2664_0_0 | - |
| 2664 | reducing_amount_of_alcohol1: Illness vs. Doctor's advice | - |
| 2664 | reducing_amount_of_alcohol2: Illness vs. Health precaution | - |
| 2664 | reducing_amount_of_alcohol3: Illness vs. Financial reasons | - |
| 2664 | reducing_amount_of_alcohol4: Illness vs. Other reason | - |
| 2867 | age_started_smoking_in_former_smokers_f2867_0_0 | - |
| 2877 | type_of_tobacco_previously_smoked_f2877_0_0 | - |
| 2877 | tobacco_previously1: None vs. Manufactured cigarettes | - |
| 2877 | tobacco_previously2: None vs. Hand-rolled cigarettes | - |
| 2877 | tobacco_previously3: None vs. Cigars or pipes | - |
| 2887 | number_of_cigarettes_previously_smoked_daily_f2887_0_0 | - |
| 2897 | age_stopped_smoking_f2897_0_0 | - |
| 2907 | ever_stopped_smoking_for_6_months_f2907_0_0 | - |
| 2926 | number_of_unsuccessful_stopsmoking_attempts_f2926_0_0 | - |
| 2936 | likelihood_of_resuming_smoking_f2936_0_0: No vs. Yes | - |
| 2966 | age_high_blood_pressure_diagnosed_f2966_0_0 | - |
| 2976 | age_diabetes_diagnosed_f2976_0_0 | - |
| 3160 | weight_manual_entry_f3160_0_0 | - |
| 3436 | age_started_smoking_in_current_smokers_f3436_0_0 | - |
| 3446 | type_of_tobacco_currently_smoked_f3446_0_0 | - |
| 3446 | tobacco_current1: None vs. Manufactured cigarettes | - |
| 3446 | tobacco_current2: None vs. Hand-rolled cigarettes | - |
| 3446 | tobacco_current3: None vs. Cigars or pipes | - |
| 3456 | number_of_cigarettes_currently_smoked_daily_current_cigarette_smokers_f3456_0_0 | sm_num_cig |
| 3466 | time_from_waking_to_first_cigarette_f3466_0_0 | - |
| 3476 | difficulty_not_smoking_for_1_day_f3476_0_0 | - |
| 3486 | ever_tried_to_stop_smoking_f3486_0_0: No vs. Yes | - |
| 3496 | wants_to_stop_smoking_f3496_0_0: No vs. Yes | - |
| 3506 | smoking_compared_to_10_years_previous_f3506_0_0 | - |
| 3627 | age_angina_diagnosed_f3627_0_0 | - |
| 3680 | age_when_last_ate_meat_f3680_0_0 | - |

| | | |
|---|---|---|
| 3731 | former_alcohol_drinker_f3731_0_0 | - |
| 3894 | age_heart_attack_diagnosed_f3894_0_0 | - |
| 4041 | gestational_diabetes_only_f4041_0_0 | - |
| 4056 | age_stroke_diagnosed_f4056_0_0 | - |
| 4079 | diastolic_blood_pressure_automated_reading_f4079_0_0 | dbt |
| 4080 | systolic_blood_pressure_automated_reading_f4080_0_0 | sbt |
| 4081 | method_of_measuring_blood_pressure_f4081_0_0 | - |
| 4081 | method_of_measuring1: Not performed vs. Question not asked due to previous answers | - |
| 4081 | method_of_measuring2: Not performed vs. Direct entry | - |
| 4081 | method_of_measuring3: Not performed vs. Manual entry of electronic results | - |
| 4081 | method_of_measuring4: Not performed vs. Manual sphygmomanometer | - |
| 4407 | average_monthly_red_wine_intake_f4407_0_0 | - |
| 4418 | average_monthly_champagne_plus_white_wine_intake_f4418_0_0 | - |
| 4429 | average_monthly_beer_plus_cider_intake_f4429_0_0 | - |
| 4440 | average_monthly_spirits_intake_f4440_0_0 | - |
| 4451 | average_monthly_fortified_wine_intake_f4451_0_0 | - |
| 4462 | average_monthly_intake_of_other_alcoholic_drinks_f4462_0_0 | - |
| 5364 | average_weekly_intake_of_other_alcoholic_drinks_f5364_0_0 | - |
| 5959 | previously_smoked_cigarettes_on_mostall_days_f5959_0_0 | - |
| 6144 | never_eat_eggs_dairy_wheat_sugar_f6144_0_0 | - |
| 6144 | never eat eggs vs. no eggs restrictions | - |
| 6144 | never eat dairy vs. no eggs, dairy, wheat, or sugar restrictions | - |
| 6144 | never eat wheat vs. no eggs, dairy, wheat, or sugar restrictions | - |
| 6144 | never eat sugar vs. no eggs, dairy, wheat, or sugar restrictions | - |
| 6150 | vascularheart_problems_diagnosed_by_doctor_f6150_0_0 | - |
| 6150 | cvd_dx1: None vs. Heart attack | - |
| 6150 | cvd_dx2: None vs. Angina | - |
| 6150 | cvd_dx3: None vs. Stroke | - |
| 6150 | cvd_dx4: None vs. High blood pressure | - |
| 6153 | medication_for_cholesterol_blood_pressure_diabetes_or_take_exogenous_hormones_f6153_0_0 | - |
| 6177 | medication_for_cholesterol_blood_pressure_or_diabetes_f6177_0_0 | - |
| 6183 | number_of_cigarettes_previously_smoked_daily_current_cigarpipe_smokers_f6183_0_0 | - |

| | | |
|---|---|---|
| 6194 | age_stopped_smoking_cigarettes_current_cigarpipe_or_previous_cigarette_smoker_f6194_0_0 | - |
| 2143 | weight_preimaging_f12143_2_0 | - |
| 2144 | height_f12144_2_0 | - |
| 20115 | country_of_birth_nonuk_origin_f20115_0_0 | - |
| 20116 | smoking_status2: current vs. never | sm_status1 |
| 20116 | smoking_status1: previous vs. never | sm_status2 |
| 20117 | alcohol_drinker_status1: current vs. never | - |
| 20117 | alcohol_drinker_status2: previous vs. never | - |
| 20160 | ever_smoked_f20160_0_0 | sm_status2 |
| 20161 | pack_years_of_smoking_f20161_0_0 | - |
| 20162 | pack_years_adult_smoking_as_proportion_of_life_span_exposed_to_smoking_f20162_0_0 | - |
| 21000 | ethnic_background_f21000_0_0 | - |
| 21001 | body_mass_index_bmi_f21001_0_0 | bmi |
| 21002 | weight_f21002_0_0 | - |
| 21003 | age_when_attended_assessment_centre_f21003_0_0 | - |
| 21022 | age_at_recruitment_f21022_0_0 | age |
| 40000 | date_of_death_f40000_0_0 | - |
| 30690 | cholesterol_f30690_0_0 | skol |
| 30740 | glucose_f30740_0_0 | blods0 |
| 30750 | glycated_haemoglobin_hba1c_f30750_0_0 | - |
| 30760 | hdl_cholesterol_f30760_0_0 | hdl |
| 30780 | ldl_direct_f30780_0_0 | ldl |
| 30870 | triglycerides_f30870_0_0 | stg |
| 34 | year_of_birth_f34_0_0 | - |
| 52 | month_of_birth_f52_0_0 | - |
| 53 | date_of_attending_assessment_centre_f53_0_0 | - |

(-): Not applicable

**Table 5. Variables removed during data processing.**

| **Trait** | **Variables** |
|---|---|
| FG (Fasting glucose) | - |
| Diastolic blood pressure (DBP) | *sm_duration.1; sm_duration.2; sbt.1; sbt.2* |

| | |
|---|---|
| 2h G (2 h glucose) | *ssn_time.1;sn_time.2 ;sm_status2.1 ;sm_status2.2* |
| High-density lipoprotein cholesterol (HDL-C) | - |
| Low-density lipoprotein cholesterol (LDL-C) | *FA160_sum1.1; FA160_sum1.2; MONOsum1.1; MONOsum1.2; mfetsum1.1; mfetsum1.2; FA140_sum1.1; FA140_sum1.2; fettsum1.1; fettsum1.2; FA170_sum1.1; FA170_sum1.2* |
| Body mass index (BMI) | *sn_time.1;sn_time.2; sm_num_cig.1; sm_num_cig.2; FA226_sum1.1; FA226_sum1.2; karosum1.1; karosum1.2; kolhsum1.1; kolhsum1.2; protsum1_anim.1; protsum1_anim.2; sm_duration.1; sm_duration.2; fettsum1.1; fettsum1.2; Folasum1.1; Folasum1.2; MONOsum1.1; MONOsum1.2 ;FA160_sum1.1; FA160_sum1.2; kolesum1.1* |
| Systolic blood pressure (SBP) | *dbt.1; dbt.2* |
| Total cholesterol (TC) | *FA160_sum1.1;FA160_sum1.2; FA226_sum1.1; FA226_sum1.2; karosum1.1; karosum1.2; mfetsum1.1; mfetsum1.2; POLYsum1.1; POLYsum1.2; fettsum1.1; fettsum1.2; Folasum1.1; Folasum1.2; MONOsum1.1; MONOsum1.2; NATRsum1.1; NATRsum1.2; NIACsum1.1; NIACsum1.2; sm_duration.1; sm_duration.2* |
| Triglycerides (TG) | *sn_time.1; sn_time.2; karosum1.1; karosum1.2; MONOsum1.1; MONOsum1.2; sm_cig_groups.1; sm_cig_groups.2; sm_duration.1; sm_duration.2; FA226_sum1.1; FA226_sum1.2* |
| (-): Not applicable | |

**Table 6. 'No persistence' category hazard ratios (HR) and 95% CIs in VHU cohort**

| | | **CVD** | | | | **CVD-mortality** | | | | **Diabetes** | | | |
|---|---|---|---|---|---|---|---|---|---|---|---|---|---|
| **Adjustment** | **Trait/Categories** | **HR** | **95% (CIs)** | | ***p*** | **HR** | **95% (CIs)** | | ***p*** | **HR** | **95% (CIs)** | | ***p*** |
| | **Fasting glucose** | | | | | | | | | | | | |
| | Ref (Expected) | 1.00 | | | | 1.00 | | | | 1.00 | | | |
| **Basic** | No persistence | 1.29 | 0.93 | 1.77 | 0.12 | 1.78 | 0.63 | 5.06 | 0.28 | 1.59 | 1.02 | 2.47 | 0.04 |
| **Partial** | No persistence | 1.22 | 0.88 | 1.68 | 0.23 | 1.74 | 0.61 | 4.94 | 0.30 | 1.55 | 0.99 | 2.42 | 0.05 |
| **Full** | No persistence | 1.21 | 0.88 | 1.67 | 0.25 | 1.65 | 0.58 | 4.69 | 0.35 | 1.55 | 0.99 | 2.42 | 0.05 |
| | **2-h Glucose** | | | | | | | | | | | | |
| | Ref (Expected) | 1.00 | | | | 1.00 | | | | 1.00 | | | |
| **Basic** | No persistence | 1.18 | 0.84 | 1.65 | 0.33 | 2.85 | 1.16 | 7.03 | 0.02 | 2.63 | 1.64 | 4.20 | 5.58E-05 |
| **Partial** | No persistence | 1.13 | 0.81 | 1.58 | 0.47 | 2.90 | 1.17 | 7.17 | 0.02 | 2.60 | 1.62 | 4.18 | 7.02E-05 |

| Adjustment | Trait/Categories | CVD HR | CVD 95% (CIs) | | CVD *p* | CVD-mortality HR | CVD-mortality 95% (CIs) | | CVD-mortality *p* | Diabetes HR | Diabetes 95% (CIs) | | Diabetes *p* |
|---|---|---|---|---|---|---|---|---|---|---|---|---|---|
| **Full** | No persistence | 1.11 | 0.79 | 1.55 | 0.54 | 2.85 | 1.15 | 7.07 | 0.02 | 2.59 | 1.61 | 4.15 | 7.91E-05 |
| | **DBP** | | | | | | | | | | | | |
| | Ref (Expected) | 1.00 | | | | 1.00 | | | | 1.00 | | | |
| **Basic** | No persistence | 1.40 | 1.03 | 1.91 | 0.03 | 2.65 | 1.03 | 6.81 | 0.04 | 0.90 | 0.46 | 1.75 | 0.75 |
| **Partial** | No persistence | 1.42 | 1.05 | 1.94 | 0.03 | 2.66 | 1.04 | 6.84 | 0.04 | 0.86 | 0.44 | 1.69 | 0.67 |
| **Full** | No persistence | 1.41 | 1.04 | 1.93 | 0.03 | 2.62 | 1.02 | 6.75 | 0.05 | 0.86 | 0.44 | 1.68 | 0.65 |
| | **HDL** | | | | | | | | | | | | |
| | Ref (Expected) | 1.00 | | | | 1.00 | | | | 1.00 | | | |
| **Basic** | No persistence | 0.48 | 0.18 | 1.31 | 0.15 | 2.07 | 0.25 | 17.47 | 0.50 | 1.52 | 0.53 | 4.34 | 0.43 |
| **Partial** | No persistence | 0.46 | 0.17 | 1.25 | 0.13 | 1.68 | 0.20 | 14.25 | 0.63 | 1.53 | 0.53 | 4.38 | 0.43 |
| **Full** | No persistence | 0.44 | 0.16 | 1.20 | 0.11 | 1.70 | 0.20 | 14.55 | 0.63 | 1.55 | 0.54 | 4.45 | 0.41 |
| | **BMI** | | | | | | | | | | | | |
| | Ref (Expected) | 1.00 | | | | 1.00 | | | | 1.00 | | | |
| **Basic** | No persistence | 0.91 | 0.61 | 1.36 | 0.65 | 0.34 | 0.04 | 2.63 | 0.30 | 1.21 | 0.75 | 1.95 | 0.43 |
| **Partial** | No persistence | 0.88 | 0.59 | 1.32 | 0.55 | 0.32 | 0.04 | 2.51 | 0.28 | 1.24 | 0.77 | 2.00 | 0.38 |
| **Full** | No persistence | 0.88 | 0.59 | 1.32 | 0.53 | 0.32 | 0.04 | 2.49 | 0.28 | 1.28 | 0.79 | 2.05 | 0.31 |
| | **LDL** | | | | | | | | | | | | |
| | Ref (Expected) | 1.00 | | | | 1.00 | | | | 1.00 | | | |
| **Basic** | No persistence | 2.24 | 1.28 | 3.91 | 4.47E-03 | 2.92 | 0.60 | 14.16 | 0.18 | 0.84 | 0.26 | 2.73 | 0.77 |
| **Partial** | No persistence | 2.23 | 1.28 | 3.90 | 4.89E-03 | 2.97 | 0.60 | 14.64 | 0.18 | 0.84 | 0.26 | 2.73 | 0.77 |
| **Full** | No persistence | 2.22 | 1.27 | 3.87 | 0.01 | 2.91 | 0.58 | 14.44 | 0.19 | 0.83 | 0.25 | 2.71 | 0.75 |
| | **Total Cholesterol** | | | | | | | | | | | | |
| | Ref (Expected) | 1.00 | | | | 1.00 | | | | 1.00 | | | |
| **Basic** | No persistence | 1.28 | 0.93 | 1.76 | 0.13 | 1.82 | 0.64 | 5.21 | 0.27 | 1.02 | 0.57 | 1.84 | 0.94 |
| **Partial** | No persistence | 1.25 | 0.91 | 1.73 | 0.17 | 1.83 | 0.64 | 5.26 | 0.26 | 1.05 | 0.58 | 1.89 | 0.87 |
| **Full** | No persistence | 1.24 | 0.90 | 1.72 | 0.18 | 1.80 | 0.63 | 5.18 | 0.27 | 1.04 | 0.58 | 1.88 | 0.89 |

| Adjustment | Trait/Categories | CVD HR | CVD 95% (CIs) | | CVD *p* | CVD-mortality HR | CVD-mortality 95% (CIs) | | CVD-mortality *p* | Diabetes HR | Diabetes 95% (CIs) | | Diabetes *p* |
|---|---|---|---|---|---|---|---|---|---|---|---|---|---|
| | **Triglycerides** | | | | | | | | | | | | |
| | Ref (Expected) | 1.00 | | | | 1.00 | | | | 1.00 | | | |
| **Basic** | No persistence | 1.15 | 0.66 | 2.01 | 0.62 | 1.89 | 0.45 | 7.94 | 0.39 | 1.67 | 0.85 | 3.27 | 0.14 |
| **Partial** | No persistence | 1.13 | 0.65 | 1.97 | 0.67 | 1.81 | 0.43 | 7.63 | 0.42 | 1.63 | 0.83 | 3.20 | 0.16 |
| **Full** | No persistence | 1.10 | 0.63 | 1.93 | 0.73 | 1.73 | 0.41 | 7.31 | 0.45 | 1.59 | 0.81 | 3.13 | 0.18 |
| | **SBP** | | | | | | | | | | | | |
| | Ref (Expected) | 1.00 | | | | 1.00 | | | | 1.00 | | | |
| **Basic** | No persistence | 1.32 | 0.96 | 1.82 | 0.09 | 1.00 | 0.24 | 4.24 | 1.00 | 0.78 | 0.43 | 1.44 | 0.43 |
| **Partial** | No persistence | 1.29 | 0.94 | 1.78 | 0.11 | 1.03 | 0.24 | 4.37 | 0.97 | 0.79 | 0.43 | 1.45 | 0.44 |
| **Full** | No persistence | 1.29 | 0.94 | 1.77 | 0.12 | 1.03 | 0.24 | 4.38 | 0.96 | 0.78 | 0.42 | 1.44 | 0.43 |

BMI: Body Mass index; SBP: systolic blood pressure; DBP: diastolic blood pressure; HDL-C: High-density lipoprotein cholesterol; LDL-C: low-density lipoprotein cholesterol.

**Table 7. Hazard ratios (HR) and 95%CI of prediction interval categories and CVD**

| | Categories | N | Events | person-years | IRR* | 95% (CIs) | | IRD* | 95% (CIs) | | HR | 95% (CIs) | | p |
|---|---|---|---|---|---|---|---|---|---|---|---|---|---|---|
| | **Fasting glucose** | | | | | | | | | | | | | |
| | Ref (Expected) | 13596 | 446 | 10854.5 | | | | | | | 1.00 | | | |
| **Basic** | Sensitive | 576 | 26 | 486.5 | 1.30 | 0.88 | 1.93 | 0.01 | -0.01 | 0.03 | 1.51 | 0.83 | 2.74 | 0.18 |
| **Basic** | Resilient | 261 | 11 | 193.1 | 1.39 | 0.76 | 2.52 | 0.02 | -0.02 | 0.05 | 1.26 | 0.85 | 1.88 | 0.24 |
| **Partial** | Sensitive | | | | | | | | | | 1.53 | 0.84 | 2.80 | 0.16 |
| **Partial** | Resilient | | | | | | | | | | 1.20 | 0.81 | 1.78 | 0.37 |
| **Full** | Sensitive | | | | | | | | | | 1.54 | 0.85 | 2.81 | 0.16 |
| **Full** | Resilient | | | | | | | | | | 1.21 | 0.82 | 1.80 | 0.34 |
| **Sensitivity** | Sensitive | | | | | | | | | | 1.54 | 0.85 | 2.81 | 0.16 |

| | Categories | N | Events | person-years | IRR* | 95% (CIs) | | IRD* | 95% (CIs) | | HR | 95% (CIs) | | p |
|---|---|---|---|---|---|---|---|---|---|---|---|---|---|---|
| **Sensitivity** | Resilient | | | | | | | | | | 1.21 | 0.82 | 1.80 | 0.34 |
| | **2-h Glucose** | | | | | | | | | | | | | |
| | Ref (Expected) | 12233 | 392 | 10063.7 | | | | | | | 1.00 | | | |
| **Basic** | Resilient | 535 | 16 | 457.9 | 0.90 | 0.54 | 1.48 | 0.00 | -0.02 | 0.01 | 0.79 | 0.48 | 1.30 | 0.35 |
| **Basic** | Sensitive | 192 | 6 | 160.3 | 0.96 | 0.43 | 2.15 | 0.00 | -0.03 | 0.03 | 1.32 | 0.59 | 2.97 | 0.50 |
| **Partial** | Resilient | | | | | | | | | | 0.77 | 0.47 | 1.27 | 0.31 |
| **Partial** | Sensitive | | | | | | | | | | 1.28 | 0.57 | 2.87 | 0.55 |
| **Full** | Resilient | | | | | | | | | | 0.77 | 0.47 | 1.27 | 0.31 |
| **Full** | Sensitive | | | | | | | | | | 1.33 | 0.59 | 2.98 | 0.49 |
| **Sensitivity** | Resilient | | | | | | | | | | 0.77 | 0.47 | 1.27 | 0.31 |
| **Sensitivity** | Sensitive | | | | | | | | | | 1.33 | 0.59 | 2.98 | 0.49 |
| | **DBP** | | | | | | | | | | | | | |
| | Ref (Expected) | 13127 | 429 | 10873.5 | | | | | | | 1.00 | | | |
| **Basic** | Resilient | 456 | 6 | 374.1 | 0.41 | 0.18 | 0.91 | -0.02 | -0.04 | -0.01 | 0.39 | 0.17 | 0.87 | 0.02 |
| **Basic** | Sensitive | 252 | 19 | 202.6 | 2.38 | 1.50 | 3.76 | 0.05 | 0.01 | 0.10 | 2.25 | 1.49 | 3.39 | 1.09E-04 |
| **Partial** | Resilient | | | | | | | | | | 0.39 | 0.17 | 0.86 | 0.02 |
| **Partial** | Sensitive | | | | | | | | | | 2.24 | 1.49 | 3.38 | 1.19E-04 |
| **Full** | Resilient | | | | | | | | | | 0.39 | 0.17 | 0.87 | 0.02 |
| **Full** | Sensitive | | | | | | | | | | 2.25 | 1.49 | 3.40 | 1.05E-04 |
| **Sensitivity** | Resilient | | | | | | | | | | 0.39 | 0.17 | 0.87 | 2.10E-02 |
| **Sensitivity** | Sensitive | | | | | | | | | | 2.25 | 1.49 | 3.40 | 1.05E-04 |
| | **HDL** | | | | | | | | | | | | | |

| | Categories | N | Events | person-years | IRR* | 95% (CIs) | | IRD* | 95% (CIs) | | HR | 95% (CIs) | | p |
|---|---|---|---|---|---|---|---|---|---|---|---|---|---|---|
| | Ref (Expected) | 1855 | 90 | 1416.9 | | | | | | | 1.00 | | | |
| **Basic** | Resilient | 95 | 3 | 70.5 | 0.67 | 0.21 | 2.12 | -0.02 | -0.07 | 0.03 | 0.64 | 0.20 | 2.02 | 0.45 |
| **Basic** | Sensitive | 91 | 5 | 66.9 | 1.18 | 0.48 | 2.89 | 0.01 | -0.06 | 0.08 | 1.13 | 0.46 | 2.78 | 0.79 |
| **Partial** | Resilient | | | | | | | | | | 0.65 | 0.21 | 2.07 | 0.47 |
| **Partial** | Sensitive | | | | | | | | | | 1.20 | 0.48 | 2.97 | 0.69 |
| **Full** | Resilient | | | | | | | | | | 0.64 | 0.20 | 2.03 | 0.45 |
| **Full** | Sensitive | | | | | | | | | | 1.22 | 0.49 | 3.01 | 0.67 |
| **Sensitivity** | Resilient | | | | | | | | | | 0.64 | 0.20 | 2.03 | 0.45 |
| **Sensitivity** | Sensitive | | | | | | | | | | 1.22 | 0.49 | 3.01 | 0.67 |
| | **BMI** | | | | | | | | | | | | | |
| | Ref (Expected) | 12951 | 423 | 10640.4 | | | | | | | 1.00 | | | |
| **Basic** | Resilient | 636 | 19 | 557.1 | 0.86 | 0.54 | 1.36 | -0.01 | -0.02 | 0.01 | 1.24 | 0.76 | 2.03 | 0.39 |
| **Basic** | Sensitive | 257 | 5 | 198.2 | 0.63 | 0.26 | 1.53 | -0.01 | -0.04 | 0.01 | 0.51 | 0.20 | 1.33 | 0.17 |
| **Partial** | Resilient | | | | | | | | | | 1.24 | 0.76 | 2.04 | 0.39 |
| **Partial** | Sensitive | | | | | | | | | | 0.51 | 0.20 | 1.32 | 0.17 |
| **Full** | Resilient | | | | | | | | | | 1.18 | 0.72 | 1.94 | 0.50 |
| **Full** | Sensitive | | | | | | | | | | 0.53 | 0.20 | 1.36 | 0.19 |
| **Sensitivity** | Resilient | | | | | | | | | | 1.18 | 0.72 | 1.94 | 0.50 |
| **Sensitivity** | Sensitive | | | | | | | | | | 0.53 | 0.20 | 1.36 | 0.19 |
| | **LDL** | | | | | | | | | | | | | |
| | Ref (Expected) | 1921 | 91 | 1447.2 | | | | | | | 1.00 | | | |
| **Basic** | Resilient | 68 | 4 | 49.8 | 1.28 | 0.47 | 3.48 | 0.02 | -0.06 | 0.10 | 1.50 | 0.55 | 4.10 | 0.43 |
| **Basic** | Sensitive | 114 | 12 | 90.8 | 2.10 | 1.15 | 3.84 | 0.07 | -0.01 | 0.15 | 2.45 | 1.33 | 4.51 | 0.00 |
| **Partial** | Resilient | | | | | | | | | | 1.48 | 0.54 | 4.05 | 0.45 |

| | Categories | N | Events | person-years | IRR* | 95% (CIs) | | IRD* | 95% (CIs) | | HR | 95% (CIs) | | p |
|---|---|---|---|---|---|---|---|---|---|---|---|---|---|---|
| **Partial** | Sensitive | | | | | | | | | | 2.38 | 1.29 | 4.40 | 0.01 |
| **Full** | Resilient | | | | | | | | | | 1.52 | 0.56 | 4.18 | 0.41 |
| **Full** | Sensitive | | | | | | | | | | 2.17 | 1.17 | 4.02 | 0.01 |
| **Sensitivity** | Resilient | | | | | | | | | | 1.52 | 0.56 | 4.18 | 0.41 |
| **Sensitivity** | Sensitive | | | | | | | | | | 2.17 | 1.17 | 4.02 | 0.01 |
| | **Total Cholesterol** | | | | | | | | | | | | | |
| | Ref (Expected) | 12637 | 403 | 10425.3 | | | | | | | 1.00 | | | |
| **Basic** | Resilient | 651 | 14 | 523.3 | 0.69 | 0.41 | 1.18 | -0.01 | -0.03 | 0.00 | 0.78 | 0.46 | 1.32 | 0.35 |
| **Basic** | Sensitive | 466 | 39 | 405.2 | 2.49 | 1.79 | 3.46 | 0.06 | 0.03 | 0.09 | 2.29 | 1.65 | 3.19 | 0.00 |
| **Partial** | Resilient | | | | | | | | | | 0.77 | 0.45 | 1.31 | 0.34 |
| **Partial** | Sensitive | | | | | | | | | | 2.25 | 1.62 | 3.13 | 0.00 |
| **Full** | Resilient | | | | | | | | | | 0.78 | 0.46 | 1.33 | 0.37 |
| **Full** | Sensitive | | | | | | | | | | 2.23 | 1.60 | 3.10 | 2.00E-06 |
| **Sensitivity** | Resilient | | | | | | | | | | 0.78 | 0.46 | 1.33 | 0.37 |
| **Sensitivity** | Sensitive | | | | | | | | | | 2.23 | 1.60 | 3.10 | 0.00 |
| | **Triglycerides** | | | | | | | | | | | | | |
| | Ref (Expected) | 10498 | 330 | 8592.6 | | | | | | | 1.00 | | | |
| **Basic** | Resilient | 1 | 0 | 1.1 | - | - | - | - | - | - | - | - | - | - |
| **Basic** | Sensitive | 378 | 16 | 311.0 | 1.34 | 0.81 | 2.21 | 0.01 | -0.01 | 0.04 | 1.12 | 0.68 | 1.86 | 0.65 |
| **Partial** | Resilient | | | | | | | | | | - | - | - | - |
| **Partial** | Sensitive | | | | | | | | | | 1.11 | 0.67 | 1.83 | 0.68 |
| **Full** | Resilient | | | | | | | | | | - | - | - | - |
| **Full** | Sensitive | | | | | | | | | | 1.11 | 0.67 | 1.83 | 0.70 |
| **Sensitivity** | Resilient | | | | | | | | | | - | - | - | - |

| | Categories | N | Events | person-years | IRR* | 95% (CIs) | | IRD* | 95% (CIs) | | HR | 95% (CIs) | | p |
|---|---|---|---|---|---|---|---|---|---|---|---|---|---|---|
| **Sensitivity** | Sensitive | | | | | | | | | | 1.11 | 0.67 | 1.83 | 0.70 |
| | **SBP** | | | | | | | | | | | | | |
| | Ref (Expected) | 13021 | 408 | 10773.1 | | | | | | | 1.00 | | | |
| **Basic** | Resilient | 474.00 | 6.00 | 377.83 | 0.71 | 0.10 | 5.05 | -0.01 | -0.06 | 0.04 | 0.45 | 0.20 | 1.02 | 0.05 |
| **Basic** | Sensitive | 201.00 | 10.00 | 162.55 | 1.66 | 0.53 | 5.18 | 0.02 | -0.05 | 0.10 | 1.59 | 0.85 | 2.98 | 0.15 |
| **Partial** | Resilient | | | | | | | | | | 0.47 | 0.21 | 1.05 | 0.06 |
| **Partial** | Sensitive | | | | | | | | | | 1.59 | 0.85 | 2.99 | 0.15 |
| **Full** | Resilient | | | | | | | | | | 0.46 | 0.21 | 1.04 | 0.06 |
| **Full** | Sensitive | | | | | | | | | | 1.63 | 0.87 | 3.05 | 0.13 |
| **Sensitivity** | Resilient | | | | | | | | | | 0.46 | 0.21 | 1.04 | 0.06 |
| **Sensitivity** | Sensitive | | | | | | | | | | 1.63 | 0.87 | 3.05 | 0.13 |

" - "  it was not possible to estimate the number.  IRR: incidence rate ratio. IRD: incidence rate differences. *Per 100,000 person-years; SBP: systolic blood pressure; DBP: diastolic blood pressure; HDL-C: High-density lipoprotein cholesterol; LDL-C: low-density lipoprotein cholesterol. Sensitivity correspond to adding two non-modififable covariates to adjust for : 'Parents or siblings have diabetes' and 'Parents or siblings had a cerebral haemorrhage/thrombosis or cardiac infarction before the age of 60'

**Table 8. Hazard ratios (HR) and 95%CI of prediction interval categories and CVD mortality**

| | Categories | N | Events | person-years | IRR* | 95% (CIs) | | IRD* | 95% (CIs) | | HR | 95% (CIs) | | p |
|---|---|---|---|---|---|---|---|---|---|---|---|---|---|---|
| | **Fasting glucose** | | | | | | | | | | | | | |
| | Ref (Expected) | 13596 | 31 | 10854.5 | | | | | | | 1.00 | | | |
| **Basic** | Sensitive | 576 | 2 | 486.5 | 1.44 | 0.34 | 6.01 | 0.00 | 0.00 | 0.01 | - | - | - | - |
| **Basic** | Resilient | 261 | 0 | 193.1 | - | - | - | - | - | - | 1.31 | 0.31 | 5.50 | 0.71 |
| **Partial** | Sensitive | | | | | | | | | | - | - | - | - |

| | Categories | N | Events | person-years | IRR* | 95% (CIs) | | IRD* | 95% (CIs) | | HR | 95% (CIs) | | p |
|---|---|---|---|---|---|---|---|---|---|---|---|---|---|---|
| **Partial** | Resilient | | | | | | | | | | 1.29 | 0.31 | 5.40 | 0.73 |
| **Full** | Sensitive | | | | | | | | | | - | - | - | - |
| **Full** | Resilient | | | | | | | | | | 1.31 | 0.31 | 5.51 | 0.71 |
| **Sensitivity** | Sensitive | | | | | | | | | | - | - | - | - |
| **Sensitivity** | Resilient | | | | | | | | | | 1.31 | 0.31 | 5.51 | 0.71 |
| | **2-h Glucose** | | | | | | | | | | | | | |
| | Ref (Expected) | 12233 | 23 | 10063.7 | | | | | | | 1.00 | | | |
| **Basic** | Resilient | 535 | 0 | 457.9 | - | - | - | - | - | - | - | - | - | - |
| **Basic** | Sensitive | 192 | 0 | 160.3 | - | - | - | - | - | - | - | - | - | - |
| **Partial** | Resilient | | | | | | | | | | - | - | - | - |
| **Partial** | Sensitive | | | | | | | | | | - | - | - | - |
| **Full** | Resilient | | | | | | | | | | - | - | - | - |
| **Full** | Sensitive | | | | | | | | | | - | - | - | - |
| **Sensitivity** | Resilient | | | | | | | | | | - | - | - | - |
| **Sensitivity** | Sensitive | | | | | | | | | | - | - | - | - |
| | **DBP** | | | | | | | | | | | | | |
| | Ref (Expected) | 13127 | 33 | 10873.5 | | | | | | | 1.00 | | | |
| **Basic** | Resilient | 456 | 0 | 374.1 | - | - | - | - | - | - | - | - | - | - |
| **Basic** | Sensitive | 252 | 1 | 202.6 | 1.63 | 0.22 | ### | 0.00 | -0.01 | 0.01 | 1.38 | 0.19 | ### | 0.75 |
| **Partial** | Resilient | | | | | | | | | | - | - | - | - |
| **Partial** | Sensitive | | | | | | | | | | 1.36 | 0.19 | 9.95 | 0.76 |
| **Full** | Resilient | | | | | | | | | | - | - | - | - |
| **Full** | Sensitive | | | | | | | | | | 1.36 | 0.19 | 9.99 | 0.76 |
| **Sensitivity** | Resilient | | | | | | | | | | - | - | - | - |

| | Categories | N | Events | person-years | IRR* | 95% (CIs) | | IRD* | 95% (CIs) | | HR | 95% (CIs) | | p |
|---|---|---|---|---|---|---|---|---|---|---|---|---|---|---|
| **Sensitivity** | Sensitive | | | | | | | | | | 1.36 | 0.19 | 9.99 | 0.76 |
| | **HDL** | | | | | | | | | | | | | |
| | Ref (Expected) | 1855 | 6 | 1416.9 | 3.53 | 0.42 | ### | 0.01 | -0.02 | 0.04 | 1.00 | | | |
| **Basic** | Resilient | 95 | 0 | 70.5 | - | - | - | - | - | - | - | - | - | - |
| **Basic** | Sensitive | 91 | 1 | 66.9 | | | | | | | 3.99 | 0.47 | ### | 0.21 |
| **Partial** | Resilient | | | | | | | | | | - | - | - | - |
| **Partial** | Sensitive | | | | | | | | | | 6.07 | 0.66 | ### | 0.11 |
| **Full** | Resilient | | | | | | | | | | - | - | - | - |
| **Full** | Sensitive | | | | | | | | | | 4.68 | 0.49 | ### | 0.18 |
| **Sensitivity** | Resilient | | | | | | | | | | - | - | - | - |
| **Sensitivity** | Sensitive | | | | | | | | | | 4.68 | 0.49 | ### | 0.18 |
| | **BMI** | | | | | | | | | | | | | |
| | Ref (Expected) | 13028 | 30 | ###### | | | | | | | 1.00 | | | |
| **Basic** | Resilient | 638 | 0 | 13024.7 | - | - | - | - | - | - | - | - | - | - |
| **Basic** | Sensitive | 258 | 2 | 4940.0 | 3.47 | 0.83 | ### | 0.00 | 0.00 | 0.00 | 0.97 | 0.13 | 7.29 | 0.98 |
| **Partial** | Resilient | | | | | | | | | | - | - | - | - |
| **Partial** | Sensitive | | | | | | | | | | 0.93 | 0.12 | 7.01 | 0.95 |
| **Full** | Resilient | | | | | | | | | | - | - | - | - |
| **Full** | Sensitive | | | | | | | | | | 0.93 | 0.12 | 7.01 | 0.95 |
| **Sensitivity** | Resilient | | | | | | | | | | - | - | - | - |
| **Sensitivity** | Sensitive | | | | | | | | | | 0.93 | 0.12 | 7.01 | 0.95 |
| | **LDL** | | | | | | | | | | | | | |
| | Ref (Expected) | 1921 | 7 | 1447.2 | | | | | | | 1.00 | | | |

| | Categories | N | Events | person-years | IRR* | 95% (CIs) | | IRD* | 95% (CIs) | | HR | 95% (CIs) | | p |
|---|---|---|---|---|---|---|---|---|---|---|---|---|---|---|
| **Basic** | Resilient | 68 | 1 | 49.8 | 4.15 | 0.51 | ### | 0.02 | -0.02 | 0.05 | 3.73 | 0.46 | ### | 0.22 |
| **Basic** | Sensitive | 114 | 2 | 90.8 | 4.55 | 0.95 | ### | 0.02 | -0.01 | 0.05 | 4.81 | 0.99 | ### | 0.05 |
| **Partial** | Resilient | | | | | | | | | | 2.88 | 0.34 | ### | 0.33 |
| **Partial** | Sensitive | | | | | | | | | | 4.34 | 0.89 | ### | 0.07 |
| **Full** | Resilient | | | | | | | | | | 2.92 | 0.33 | ### | 0.34 |
| **Full** | Sensitive | | | | | | | | | | 3.99 | 0.81 | ### | 0.09 |
| **Sensitivity** | Resilient | | | | | | | | | | 2.92 | 0.33 | ### | 0.34 |
| **Sensitivity** | Sensitive | | | | | | | | | | 3.99 | 0.81 | ### | 0.09 |
| | **Total Cholesterol** | | | | | | | | | | | | | |
| | Ref (Expected) | 12637 | 27 | 10425.3 | | | | | | | 1.00 | | | |
| **Basic** | Resilient | 651 | 2 | 523.3 | 1.48 | 0.35 | 6.21 | 0.00 | 0.00 | 0.01 | 1.58 | 0.37 | 6.79 | 0.53 |
| **Basic** | Sensitive | 466 | 3 | 405.2 | 2.86 | 0.87 | 9.42 | 0.00 | 0.00 | 0.01 | 2.50 | 0.76 | 8.24 | 0.13 |
| **Partial** | Resilient | | | | | | | | | | 1.62 | 0.38 | 6.94 | 0.51 |
| **Partial** | Sensitive | | | | | | | | | | 2.40 | 0.73 | 7.95 | 0.15 |
| **Full** | Resilient | | | | | | | | | | 1.74 | 0.41 | 7.40 | 0.45 |
| **Full** | Sensitive | | | | | | | | | | 2.33 | 0.70 | 7.72 | 0.17 |
| **Sensitivity** | Resilient | | | | | | | | | | 1.74 | 0.41 | 7.40 | 0.45 |
| **Sensitivity** | Sensitive | | | | | | | | | | 2.33 | 0.70 | 7.72 | 0.17 |
| | **Triglycerides** | | | | | | | | | | | | | |
| | Ref (Expected) | 10498 | 29 | 8592.6 | | | | | | | 1.00 | | | |
| **Basic** | Resilient | 1 | 0 | 1.1 | - | - | - | - | - | - | - | - | - | - |
| **Basic** | Sensitive | 378 | 2 | 311.0 | 1.91 | 0.45 | 7.99 | 0.00 | -0.01 | 0.01 | 1.52 | 0.36 | 6.40 | 0.56 |
| **Partial** | Resilient | | | | | | | | | | - | - | - | - |
| **Partial** | Sensitive | | | | | | | | | | 1.45 | 0.34 | 6.10 | 0.61 |

| | Categories | N | Events | person-years | IRR* | 95% (CIs) | | IRD* | 95% (CIs) | | HR | 95% (CIs) | | p |
|---|---|---|---|---|---|---|---|---|---|---|---|---|---|---|
| **Full** | Resilient | | | | | | | | | | - | - | - | - |
| **Full** | Sensitive | | | | | | | | | | 1.43 | 0.34 | 6.00 | 0.63 |
| **Sensitivity** | Resilient | | | | | | | | | | - | - | - | - |
| **Sensitivity** | Sensitive | | | | | | | | | | 1.43 | 0.34 | 6.00 | 0.63 |
| | **SBP** | | | | | | | | | | | | | |
| | Ref (Expected) | 13021 | 25 | 10773.1 | | | | | | | 1.00 | | | |
| **Basic** | Resilient | 474.00 | 0.00 | 377.83 | - | - | - | - | - | - | - | - | - | - |
| **Basic** | Sensitive | 201.00 | 1.00 | 162.55 | 2.65 | 0.36 | ### | 0.00 | -0.01 | 0.02 | 2.42 | 0.33 | ### | 0.39 |
| **Partial** | Resilient | | | | | | | | | | - | - | - | - |
| **Partial** | Sensitive | | | | | | | | | | 2.38 | 0.32 | ### | 0.40 |
| **Full** | Resilient | | | | | | | | | | - | - | - | - |
| **Full** | Sensitive | | | | | | | | | | 2.48 | 0.33 | ### | 0.37 |
| **Sensitivity** | Resilient | | | | | | | | | | - | - | - | - |
| **Sensitivity** | Sensitive | | | | | | | | | | 2.48 | 0.33 | ### | 0.37 |

" - "  it was not possible to estimate the number.  IRR: incidence rate ratio. IRD: incidence rate differences. *Per 100,000 person-years; SBP: systolic blood pressure; DBP: diastolic blood pressure; HDL-C: High-density lipoprotein cholesterol; LDL-C: low-density lipoprotein cholesterol. Sensitivity correspond to adding two non-modififable covariates to adjust for : 'Parents or siblings have diabetes' and 'Parents or siblings had a cerebral haemorrhage/thrombosis or cardiac infarction before the age of 60'

**Table 9. Hazard ratios (HR) and 95%CI of prediction interval categories and diabetes**

| | Categories | N | Events | person-years | IRR* | 95% (CIs) | | IRD* | 95% (CIs) | | HR | 95% (CIs) | | p |
|---|---|---|---|---|---|---|---|---|---|---|---|---|---|---|
| | **Fasting glucose** | | | | | | | | | | | | | |
| | Ref (Expected) | 13566 | 186 | 10838.4 | | | | | | | 1.00 | | | |

| | Categories | N | Events | person-years | IRR* | 95% (CIs) | | IRD* | 95% (CIs) | | HR | 95% (CIs) | | p |
|---|---|---|---|---|---|---|---|---|---|---|---|---|---|---|
| **Basic** | Sensitive | 574 | 8 | 485.1 | 0.96 | 0.47 | 1.95 | 0.00 | -0.01 | 0.01 | 0.74 | 0.18 | 2.98 | 0.67 |
| **Basic** | Resilient | 261 | 2 | 193.1 | 0.60 | 0.15 | 2.43 | -0.01 | -0.02 | 0.01 | 0.85 | 0.42 | 1.72 | 0.64 |
| **Partial** | Sensitive | | | | | | | | | | 0.73 | 0.18 | 2.94 | 0.65 |
| **Partial** | Resilient | | | | | | | | | | 0.82 | 0.41 | 1.67 | 0.59 |
| **Full** | Sensitive | | | | | | | | | | 0.73 | 0.18 | 2.97 | 0.67 |
| **Full** | Resilient | | | | | | | | | | 0.83 | 0.41 | 1.68 | 0.60 |
| **Sensitivity** | Sensitive | | | | | | | | | | 0.73 | 0.18 | 2.97 | 0.67 |
| **Sensitivity** | Resilient | | | | | | | | | | 0.83 | 0.41 | 1.68 | 0.60 |
| | **2-h Glucose** | | | | | | | | | | | | | |
| | Ref (Expected) | 12210 | 110 | 10049.4 | | | | | | | 1.00 | | | |
| **Basic** | Resilient | 535 | 8 | 457.9 | 1.60 | 0.78 | 3.27 | 0.01 | -0.01 | 0.02 | 1.39 | 0.68 | 2.85 | 0.37 |
| **Basic** | Sensitive | 192 | 2 | 160.3 | 1.14 | 0.28 | 4.61 | 0.00 | -0.02 | 0.02 | 1.51 | 0.37 | 6.12 | 0.57 |
| **Partial** | Resilient | | | | | | | | | | 1.42 | 0.69 | 2.91 | 0.34 |
| **Partial** | Sensitive | | | | | | | | | | 1.47 | 0.36 | 5.97 | 0.59 |
| **Full** | Resilient | | | | | | | | | | 1.42 | 0.69 | 2.91 | 0.34 |
| **Full** | Sensitive | | | | | | | | | | 1.46 | 0.36 | 5.94 | 0.59 |
| **Sensitivity** | Resilient | | | | | | | | | | 1.42 | 0.69 | 2.91 | 0.34 |
| **Sensitivity** | Sensitive | | | | | | | | | | 1.46 | 0.36 | 5.94 | 0.59 |
| | **DBP** | | | | | | | | | | | | | |
| | Ref (Expected) | 13094 | 175 | 10851.3 | | | | | | | 1.00 | | | |
| **Basic** | Resilient | 456 | 4 | 374.1 | 0.66 | 0.25 | 1.79 | -0.01 | -0.02 | 0.01 | 0.95 | 0.39 | 2.32 | 0.92 |
| **Basic** | Sensitive | 251 | 3 | 201.7 | 0.92 | 0.29 | 2.89 | 0.00 | -0.02 | 0.02 | 0.78 | 0.25 | 2.43 | 0.66 |
| **Partial** | Resilient | | | | | | | | | | 0.94 | 0.39 | 2.29 | 0.89 |

| | Categories | N | Events | person-years | IRR* | 95% (CIs) | | IRD* | 95% (CIs) | | HR | 95% (CIs) | | p |
|---|---|---|---|---|---|---|---|---|---|---|---|---|---|---|
| **Partial** | Sensitive | | | | | | | | | | 0.78 | 0.25 | 2.46 | 0.68 |
| **Full** | Resilient | | | | | | | | | | 0.94 | 0.39 | 2.28 | 0.89 |
| **Full** | Sensitive | | | | | | | | | | 0.78 | 0.25 | 2.44 | 0.67 |
| **Sensitivity** | Resilient | | | | | | | | | | 0.94 | 0.39 | 2.28 | 0.89 |
| **Sensitivity** | Sensitive | | | | | | | | | | 0.78 | 0.25 | 2.44 | 0.67 |
| | **HDL** | | | | | | | | | | | | | |
| | Ref (Expected) | 1849 | 31 | 1413.3 | | | | | | | 1.00 | | | |
| **Basic** | Resilient | 95 | 0 | 70.5 | - | - | - | - | - | - | - | - | - | - |
| **Basic** | Sensitive | 90 | 2 | 66.1 | 1.38 | 0.33 | 5.76 | 0.01 | -0.03 | 0.05 | 1.69 | 0.40 | 7.13 | 0.47 |
| **Partial** | Resilient | | | | | | | | | | - | - | - | - |
| **Partial** | Sensitive | | | | | | | | | | 1.73 | 0.41 | 7.37 | 0.46 |
| **Full** | Resilient | | | | | | | | | | - | - | - | - |
| **Full** | Sensitive | | | | | | | | | | 1.71 | 0.40 | 7.28 | 0.47 |
| **Sensitivity** | Resilient | | | | | | | | | | - | - | - | - |
| **Sensitivity** | Sensitive | | | | | | | | | | 1.71 | 0.40 | 7.28 | 0.47 |
| | **BMI** | | | | | | | | | | | | | |
| | Ref (Expected) | 12921 | 161 | 10621.1 | | | | | | | 1.00 | | | |
| **Basic** | Resilient | 636 | 2 | 557.1 | 0.24 | 0.06 | 0.96 | -0.01 | -0.02 | -0.01 | 0.63 | 0.15 | 2.58 | 0.52 |
| **Basic** | Sensitive | 255 | 12 | 196.7 | 4.02 | 2.24 | 7.23 | 0.05 | 0.01 | 0.08 | 0.58 | 0.24 | 1.39 | 0.22 |
| **Partial** | Resilient | | | | | | | | | | 0.63 | 0.15 | 2.57 | 0.52 |
| **Partial** | Sensitive | | | | | | | | | | 0.60 | 0.25 | 1.42 | 0.25 |
| **Full** | Resilient | | | | | | | | | | 0.58 | 0.14 | 2.36 | 0.45 |
| **Full** | Sensitive | | | | | | | | | | 0.64 | 0.27 | 1.53 | 0.32 |
| **Sensitivity** | Resilient | | | | | | | | | | 0.58 | 0.14 | 2.36 | 0.45 |

| | Categories | N | Events | person-years | IRR* | 95% (CIs) | | IRD* | 95% (CIs) | | HR | 95% (CIs) | | p |
|---|---|---|---|---|---|---|---|---|---|---|---|---|---|---|
| **Sensitivity** | Sensitive | | | | | | | | | | 0.64 | 0.27 | 1.53 | 0.32 |
| | **LDL** | | | | | | | | | | | | | |
| | Ref (Expected) | 1914 | 38 | 1442.3 | | | | | | | 1.00 | | | |
| **Basic** | Resilient | 67 | 1 | 49.0 | 0.78 | 0.11 | 5.65 | -0.01 | -0.05 | 0.03 | 0.78 | 0.11 | 5.69 | 0.81 |
| **Basic** | Sensitive | 112 | 1 | 89.9 | 0.42 | 0.06 | 3.08 | -0.02 | -0.04 | 0.01 | 0.41 | 0.06 | 2.99 | 0.38 |
| **Partial** | Resilient | | | | | | | | | | 0.79 | 0.11 | 5.74 | 0.81 |
| **Partial** | Sensitive | | | | | | | | | | 0.41 | 0.06 | 2.99 | 0.38 |
| **Full** | Resilient | | | | | | | | | | 0.82 | 0.11 | 5.99 | 0.84 |
| **Full** | Sensitive | | | | | | | | | | 0.39 | 0.05 | 2.85 | 0.35 |
| **Sensitivity** | Resilient | | | | | | | | | | 0.82 | 0.11 | 5.99 | 0.84 |
| **Sensitivity** | Sensitive | | | | | | | | | | 0.39 | 0.05 | 2.85 | 0.35 |
| | **Total Cholesterol** | | | | | | | | | | | | | |
| | Ref (Expected) | 12610 | 150 | 10408.0 | | | | | | | 1.00 | | | |
| **Basic** | Resilient | 649 | 9 | 522.4 | 1.20 | 0.61 | 2.34 | 0.00 | -0.01 | 0.01 | 1.04 | 0.52 | 2.09 | 0.91 |
| **Basic** | Sensitive | 463 | 11 | 404.0 | 1.89 | 1.02 | 3.49 | 0.01 | 0.00 | 0.03 | 1.73 | 0.94 | 3.19 | 0.08 |
| **Partial** | Resilient | | | | | | | | | | 1.10 | 0.55 | 2.20 | 0.80 |
| **Partial** | Sensitive | | | | | | | | | | 1.74 | 0.94 | 3.21 | 0.08 |
| **Full** | Resilient | | | | | | | | | | 1.18 | 0.59 | 2.36 | 0.63 |
| **Full** | Sensitive | | | | | | | | | | 1.70 | 0.92 | 3.14 | 0.09 |
| **Sensitivity** | Resilient | | | | | | | | | | 1.18 | 0.59 | 2.36 | 0.63 |
| **Sensitivity** | Sensitive | | | | | | | | | | 1.70 | 0.92 | 3.14 | 0.09 |
| | **Triglycerides** | | | | | | | | | | | | | |
| | Ref (Expected) | 10469 | 148 | 8574.8 | | | | | | | 1.00 | | | |

| | Categories | N | Events | person-years | IRR* | 95% (CIs) | | IRD* | 95% (CIs) | | HR | 95% (CIs) | | p |
|---|---|---|---|---|---|---|---|---|---|---|---|---|---|---|
| **Basic** | Resilient | 1 | 0 | 1.1 | - | - | - | - | - | - | - | - | - | - |
| **Basic** | Sensitive | 376 | 4 | 309.6 | 0.75 | 0.28 | 2.02 | 0.00 | -0.02 | 0.01 | 0.69 | 0.26 | 1.87 | 0.47 |
| **Partial** | Resilient | | | | | | | | | | - | - | - | - |
| **Partial** | Sensitive | | | | | | | | | | 0.70 | 0.26 | 1.89 | 0.48 |
| **Full** | Resilient | | | | | | | | | | - | - | - | - |
| **Full** | Sensitive | | | | | | | | | | 0.69 | 0.25 | 1.86 | 0.46 |
| **Sensitivity** | Resilient | | | | | | | | | | - | - | - | - |
| **Sensitivity** | Sensitive | | | | | | | | | | 0.69 | 0.25 | 1.86 | 0.46 |
| | **SBP** | | | | | | | | | | | | | |
| | Ref (Expected) | 12996 | 170 | 10757.8 | | | | | | | 1.00 | | | |
| **Basic** | Resilient | 474.00 | 5.00 | 377.83 | 0.84 | 0.34 | 2.04 | 0.00 | -0.01 | 0.01 | 0.60 | 0.24 | 1.52 | 0.28 |
| **Basic** | Sensitive | 200.00 | 5.00 | 161.41 | 1.96 | 0.81 | 4.77 | 0.02 | -0.01 | 0.04 | 1.72 | 0.71 | 4.19 | 0.23 |
| **Partial** | Resilient | | | | | | | | | | 0.64 | 0.26 | 1.62 | 0.35 |
| **Partial** | Sensitive | | | | | | | | | | 1.74 | 0.71 | 4.23 | 0.22 |
| **Full** | Resilient | | | | | | | | | | 0.65 | 0.26 | 1.64 | 0.37 |
| **Full** | Sensitive | | | | | | | | | | 1.74 | 0.71 | 4.24 | 0.22 |
| **Sensitivity** | Resilient | | | | | | | | | | 0.65 | 0.26 | 1.64 | 0.37 |
| **Sensitivity** | Sensitive | | | | | | | | | | 1.74 | 0.71 | 4.24 | 0.22 |

" - " it was not possible to estimate the number. IRR: incidence rate ratio. IRD: incidence rate differences. *Per 100,000 person-years; SBP: systolic blood pressure; DBP: diastolic blood pressure; HDL-C: High-density lipoprotein cholesterol; LDL-C: low-density lipoprotein cholesterol. Sensitivity correspond to adding two non-modififable covariates to adjust for : 'Parents or siblings have diabetes' and 'Parents or siblings had a cerebral haemorrhage/thrombosis or cardiac infarction before the age of 60'

**Table 10. R packages used for the analyses in the current study.**

| Step | R Package |
|---|---|

| | |
|---|---|
| Variable processing | *caret [1]* |
| Missing data imputation | *missForest [2]* |
| Multicollinearity | *car [3]* |
| Performance and model assumption | *Performance [4]* |
| Quantile regression modelling | *quantreg [5]* |
| Quantile regression forest modelling | *quantregForest [6]; randomForest* |
| Cox modelling | *Survival [7]* |
| Cox proportional assumption | *Rms [8]* |
| Incidence rate ratio and | *fmsb[9]* |
| Meta-analysis | *Meta [10]* |

**Table 11. Fully-adjusted Hazard ratios and 95%CI of prediction interval categories and CVD-mortality.**

| Trait/study | Categories | n | Events | person-years | IRR* | 95% (CIs) | | IRD* | 95% (CIs) | | HR | 95% (CIs) | | *p* |
|---|---|---|---|---|---|---|---|---|---|---|---|---|---|---|
| ***Fasting glucose*** | Ref (Expected) | 13596 | 31 | 10854.5 | | | | | | | 1.00 | | | |
| **VHU** | Resilient | 261 | 0 | 193.1 | - | - | - | - | - | - | - | - | - | - |
| | Sensitive | 576 | 2 | 486.5 | 1.44 | 0.34 | 6.01 | 0.001 | -0.005 | 0.01 | 1.31 | 0.31 | 5.51 | 0.71 |
| | Ref (Expected) | 11530 | 13 | 11530 | | | | | | | | | | |
| **UKB** | Resilient | 491 | 0 | 491 | - | - | - | - | - | - | - | - | - | - |
| | Sensitive | 566 | 0 | 566 | - | - | - | - | - | - | - | - | - | - |
| ***2-h Glucose/ HbA1c*** | | | | | | | | | | | | | | |
| | Ref (Expected) | 12233 | 23 | 10063.7 | | | | | | | 1.00 | | | |
| **VHU** | Resilient | 535 | 0 | 457.9 | - | - | - | - | - | - | - | - | - | - |
| | Sensitive | 192 | 0 | 160.3 | - | - | - | - | - | - | - | - | - | - |
| | Ref (Expected) | 11493 | 7 | 262.6 | | | | | | | 1.00 | | | |
| **UKB** | Resilient | 567 | 1 | 13.1 | 2.86 | 0.35 | 23.28 | 0.05 | -0.10 | 0.20 | 3.20 | 0.39 | 26.20 | 0.28 |
| | Sensitive | 517 | 1 | 11.7 | 3.21 | 0.39 | 26.06 | 0.05 | -0.10 | 0.20 | 3.02 | 0.37 | 24.96 | 0.30 |
| ***DBP*** | | | | | | | | | | | | | | |

| Trait/study | Categories | n | Events | person-years | IRR* | 95% (CIs) | | IRD* | 95% (CIs) | | HR | 95% (CIs) | | *p* |
|---|---|---|---|---|---|---|---|---|---|---|---|---|---|---|
| | Ref (Expected) | 13127 | 33 | 10873.5 | | | | | | | 1.00 | | | |
| **VHU** | Resilient | 456 | 0 | 374.1 | - | - | - | - | - | - | - | - | - | - |
| | Sensitive | 252 | 1 | 202.6 | 1.63 | 0.22 | 11.89 | 0.002 | -0.01 | 0.01 | 1.36 | 0.19 | 9.99 | 0.76 |
| | Ref (Expected) | 11927 | 12 | 272.7 | | | | | | | 1.00 | | | |
| **UKB** | Resilient | 382 | 0 | 8.7 | - | - | - | - | - | - | - | - | - | - |
| | Sensitive | 282 | 0 | 6.5 | - | - | - | - | - | - | - | - | - | - |
| ***HDL*** | | | | | | | | | | | | | | |
| | Ref (Expected) | 1855 | 6 | 1416.9 | | | | | | | 1.00 | | | |
| **VHU** | Resilient | 95 | 0 | 70.5 | - | - | - | - | - | - | - | - | - | - |
| | Sensitive | 91 | 1 | 66.9 | 3.53 | 0.42 | 29.30 | 0.01 | -0.02 | 0.04 | 4.68 | 0.49 | 44.92 | 0.18 |
| | Ref (Expected) | 11974 | 5 | 273.8 | | | | | | | 1.00 | | | |
| **UKB** | Resilient | 288 | 0 | 6.7 | - | - | - | - | - | - | - | - | - | - |
| | Sensitive | 319 | 0 | 7.2 | - | - | - | - | - | - | - | - | - | - |
| ***BMI*** | | | | | | | | | | | | | | |
| | Ref (Expected) | 13028 | 30 | 257075.9 | | | | | | | 1.00 | | | |
| **VHU** | Resilient | 638 | 0 | 13024.7 | - | - | - | - | - | - | - | - | - | - |
| | Sensitive | 258 | 2 | 4940.0 | 3.47 | 0.83 | 14.52 | 0.0003 | -0.0003 | 0.001 | 0.93 | 0.12 | 7.01 | 0.95 |
| | Ref (Expected) | 11958 | 11 | 273.7 | | | | | | | 1.00 | | | |
| **UKB** | Resilient | 344 | 0 | 7.9 | - | - | - | - | - | - | - | - | - | - |
| | Sensitive | 271 | 0 | 6.2 | - | - | - | - | - | - | - | - | - | - |
| ***LDL*** | | | | | | | | | | | | | | |
| | Ref (Expected) | 1921 | 7 | 1447.2 | | | | | | | 1.00 | | | |
| **VHU** | Resilient | 68 | 1 | 49.8 | 4.15 | 0.51 | 33.74 | 0.02 | -0.02 | 0.05 | 2.92 | 0.33 | 25.77 | 0.34 |
| | Sensitive | 114 | 2 | 90.8 | 4.55 | 0.95 | 21.92 | 0.02 | -0.01 | 0.05 | 3.99 | 0.81 | 19.70 | 0.09 |
| | Ref (Expected) | 12422 | 10 | 284.1 | | | | | | | 1.00 | | | |
| **UKB** | Resilient | 97 | 0 | 2.2 | - | - | - | - | - | - | - | - | - | - |

| Trait/study | Categories | n | Events | person-years | IRR* | 95% (CIs) | | IRD* | 95% (CIs) | | HR | 95% (CIs) | | *p* |
|---|---|---|---|---|---|---|---|---|---|---|---|---|---|---|
| | Sensitive | 57 | 0 | 1.3 | - | - | - | - | - | - | - | - | - | - |
| ***Total Cholesterol*** | | | | | | | | | | | | | | |
| | Ref (Expected) | 12637 | 27 | 10425.3 | | | | | | | 1.00 | | | |
| **VHU** | Resilient | 651 | 2 | 523.3 | 1.48 | 0.35 | 6.21 | 0.001 | -0.004 | 0.01 | 1.74 | 0.41 | 7.40 | 0.45 |
| | Sensitive | 466 | 3 | 405.2 | 2.86 | 0.87 | 9.42 | 0.005 | -0.004 | 0.01 | 2.33 | 0.70 | 7.72 | 0.17 |
| | Ref (Expected) | 12483 | 10 | 285.3 | | | | | | | 1.00 | | | |
| **UKB** | Resilient | 34 | 0 | 0.8 | - | - | - | - | - | - | - | - | - | - |
| | Sensitive | 58 | 0 | 1.3 | - | - | - | - | - | - | - | - | - | - |
| ***Triglycerides*** | | | | | | | | | | | | | | |
| | Ref (Expected) | 10498 | 29 | 8592.6 | | | | | | | 1.00 | | | |
| **VHU** | Resilient | 1 | 0 | 1.1 | - | - | - | - | - | - | - | - | - | - |
| | Sensitive | 378 | 2 | 311.0 | 1.91 | 0.45 | 7.99 | 0.003 | -0.01 | 0.01 | 1.43 | 0.34 | 6.00 | 0.63 |
| | Ref (Expected) | 11734 | 10 | 268.5 | | | | | | | 1.00 | | | |
| **UKB** | Resilient | 465 | 0 | 10.7 | - | - | - | - | - | - | - | - | - | - |
| | Sensitive | 375 | 0 | 8.5 | - | - | - | - | - | - | - | - | - | - |
| ***SBP*** | | | | | | | | | | | | | | |
| | Ref (Expected) | 13021 | 25 | 10773.1 | | | | | | | 1.00 | | | |
| **VHU** | Resilient | 474.00 | 0.00 | 377.83 | - | - | - | - | - | - | - | - | - | - |
| | Sensitive | 201.00 | 1.00 | 162.55 | 2.65 | 0.36 | 19.57 | 0.004 | -0.01 | 0.02 | 2.48 | 0.33 | 18.46 | 0.37 |
| | Ref (Expected) | 11912 | 12 | 272.4 | | | | | | | 1.00 | | | |
| **UKB** | Resilient | 388 | 0 | 8.8 | - | - | - | - | - | - | - | - | - | - |
| | Sensitive | 291 | 0 | 6.7 | - | - | - | - | - | - | - | - | - | - |

" - "  it was not possible to estimate the number.  IRR: incidence rate ratio. IRD: incidence rate differences. *Per 100,000 person-years;  SBP: systolic blood pressure; DBP: diastolic blood pressure; HDL-C: High-density lipoprotein cholesterol; LDL-C: low-density lipoprotein cholesterol; CVD: Cardiovascular disease. T2D: Type 2 diabetes.

### SUPPLEMENTARY FIGURES

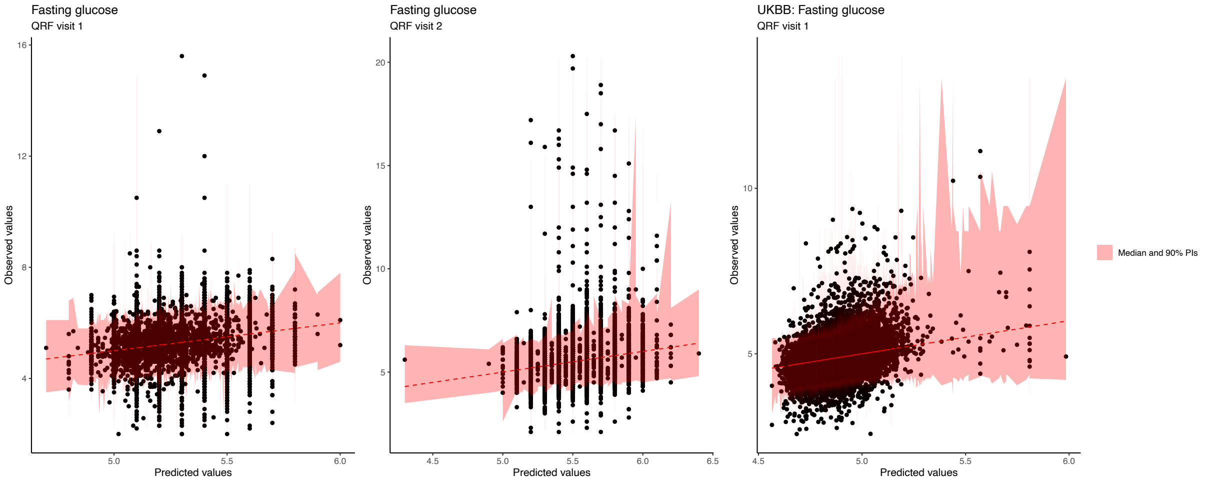

Figure 1. Scatter plots of predicted and observed values of Fasting glucose (FG) in mmol/L. In pink are depicted the 90% prediction intervals (PIs) with conditional median from quantile regression forest (QRF); From left to right, the two visits for VHU and the baseline visit for UKB.

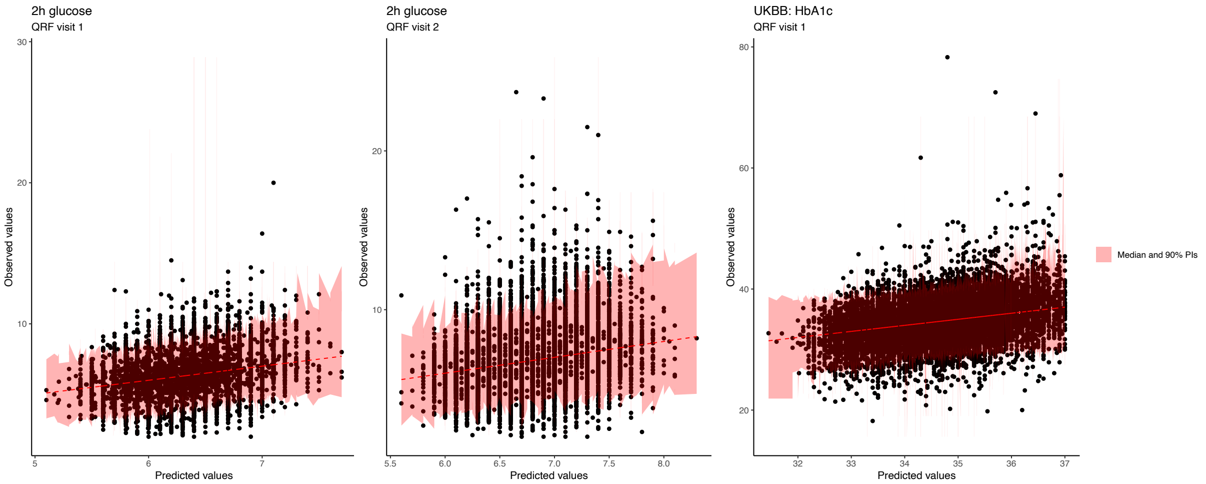

Figure 2. Scatter plots of predicted and observed values of 2-hour glucose in mmol/L and HbA1c (UKB) mmol/mol.

In pink are depicted the 90% prediction intervals (PIs) with conditional median from quantile regression forest (QRF); From left to right, the two visits for VHU and the baseline visit for UKB.

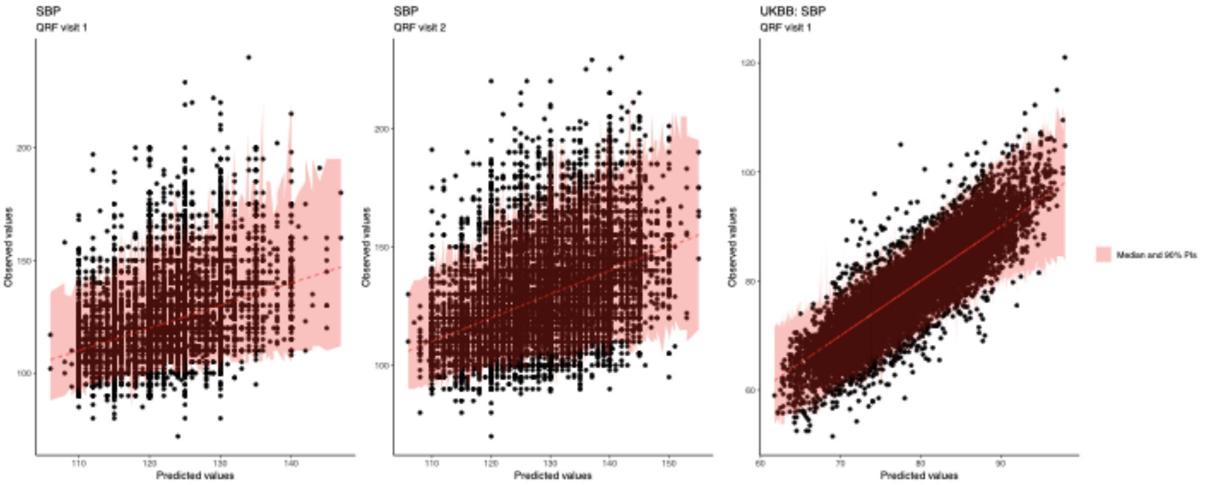

Figure 3. Scatter plots of predicted and observed values of Systolic blood pressure (SBP) in mm/Hg.
In pink are depicted the 90% prediction intervals (PIs) with conditional median from quantile regression forest (QRF); From left to right, the two visits for VHU and the baseline visit for UKB.

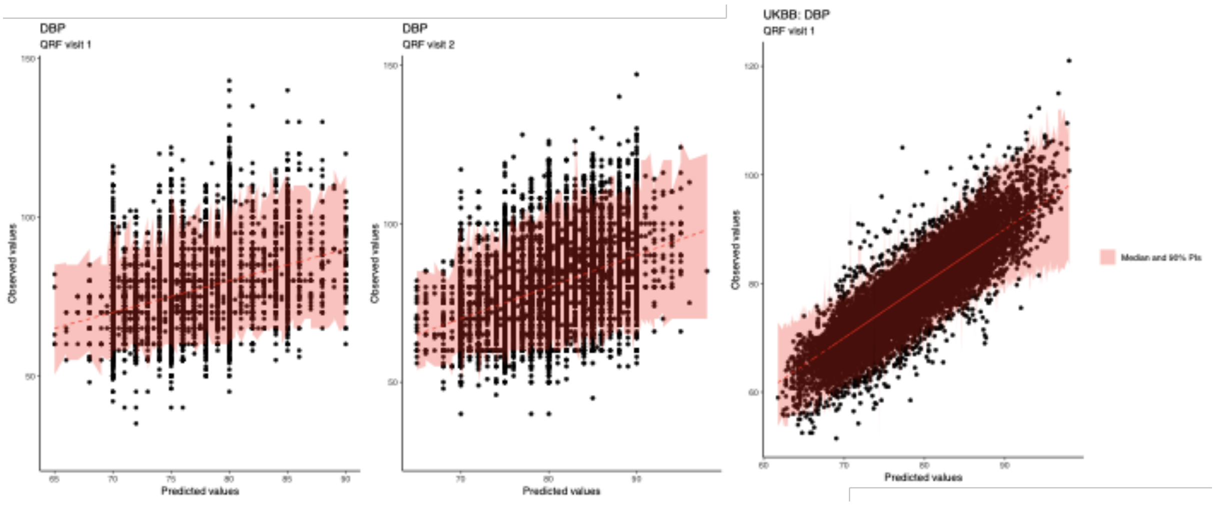

Figure 4. Scatter plots of predicted and observed values of Diastolic blood pressure (DBP) in mm/Hg.
In pink are depicted the 90% prediction intervals (PIs) with conditional median from quantile regression forest (QRF); From left to right, the two visits for VHU and the baseline visit for UKB.

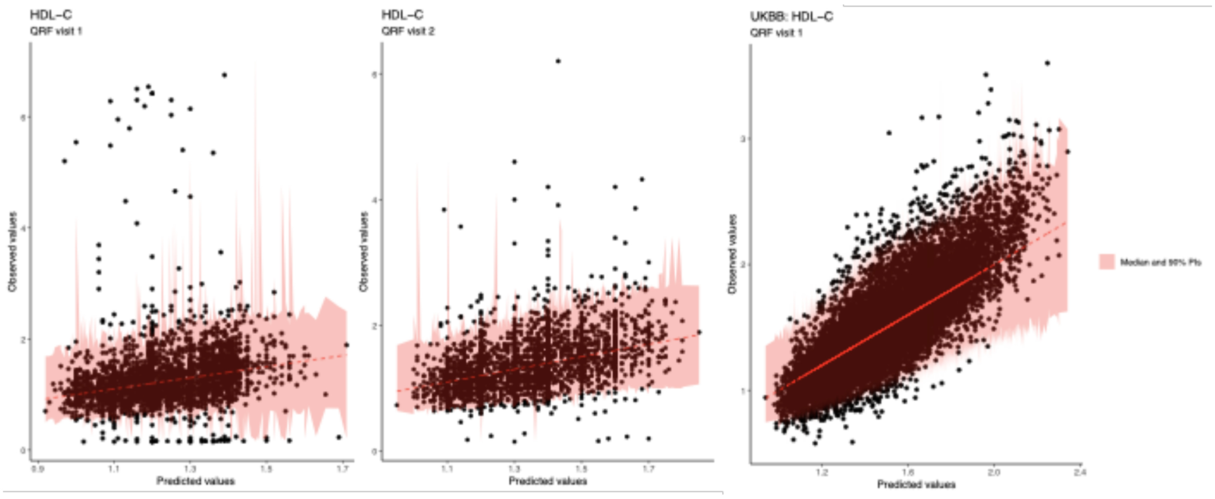

Figure 5. Scatter plots of predicted and observed values of high-density lipoprotein cholesterol (HDL-C) in mmol/L.
In pink are depicted the 90% prediction intervals (PIs) with conditional median from quantile regression forest (QRF); From left to right, the two visits for VHU and the baseline visit for UKB.

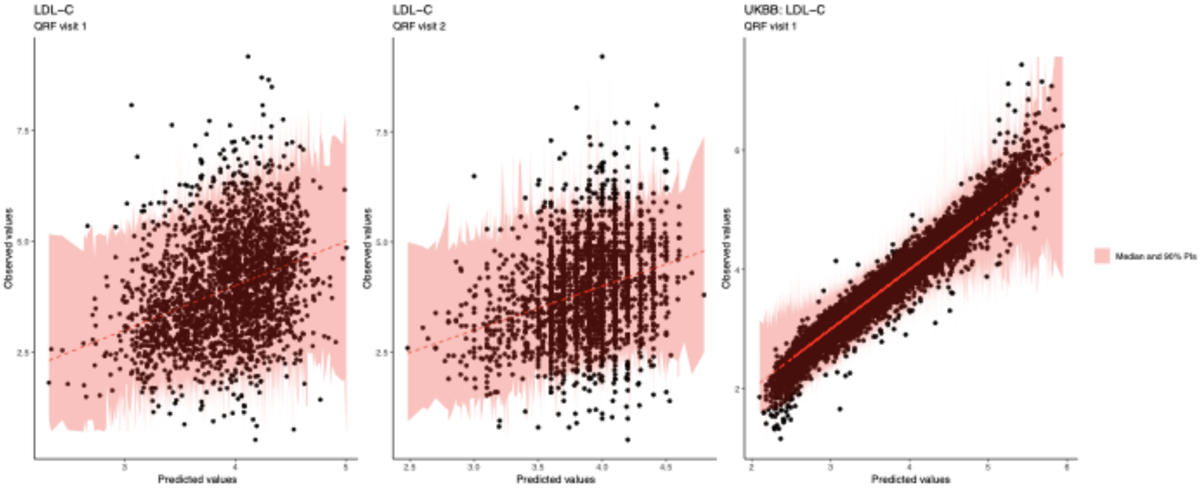

Figure 6. Scatter plots of predicted and observed values of low-density lipoprotein cholesterol (LDL-C) in mmol/L.
In pink are depicted the 90% prediction intervals (PIs) with conditional median from quantile regression forest (QRF); From left to right, the two visits for VHU and the baseline visit for UKB.

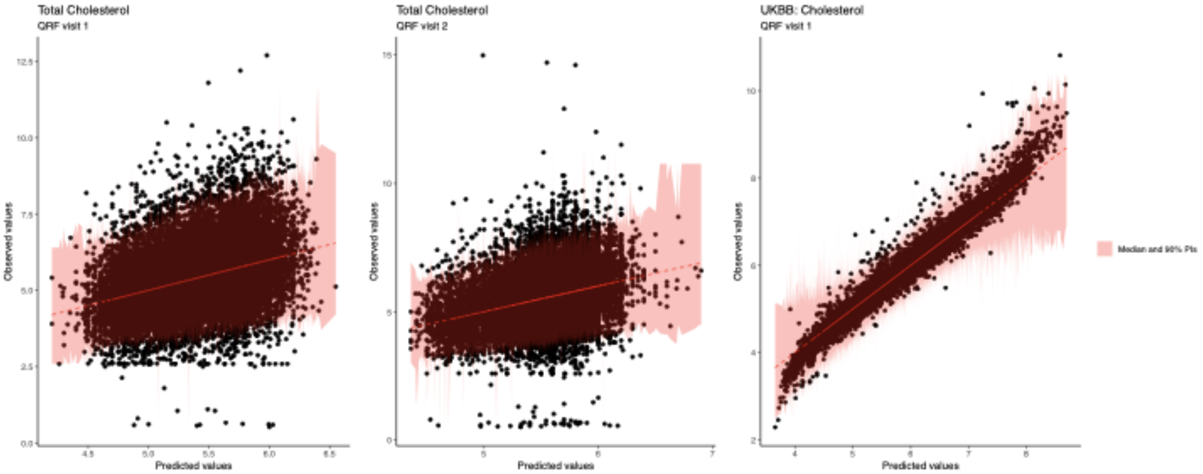

Figure 7. Scatter plots of predicted and observed values of total cholesterol (TC) in mmol/L.
In pink are depicted the 90% prediction intervals (PIs) with conditional median from quantile regression forest (QRF); From left to right, the two visits for VHU and the baseline visit for UKB.

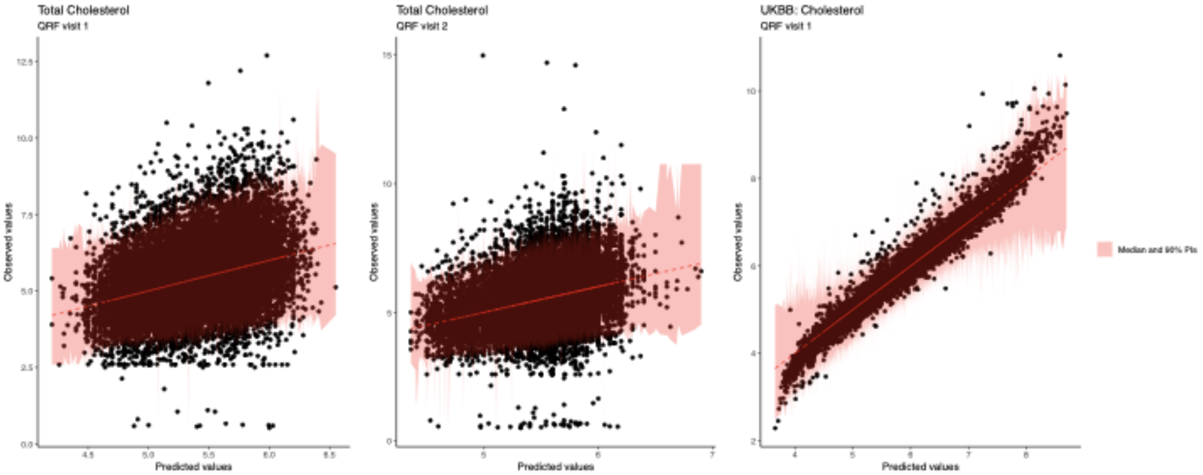

Figure 8. Scatter plots of predicted and observed values of triglycerides (TG) in mmol/L. In pink are depicted the 90% prediction intervals (PIs) with conditional median from quantile regression forest (QRF); From left to right, the two visits for VHU and the baseline visit for UKB.

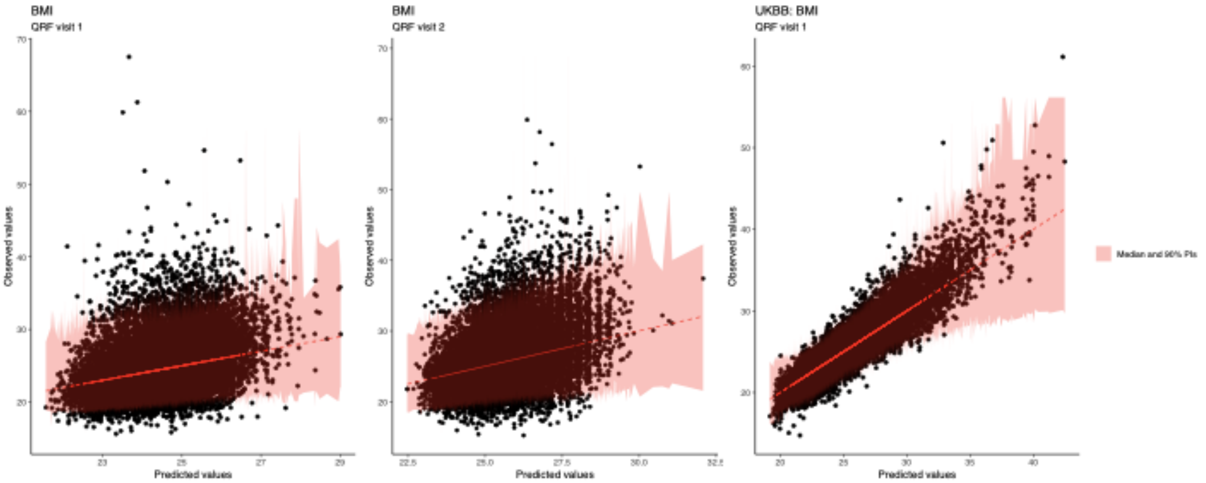

Figure 9. Scatter plots of predicted and observed values of body mass index (BMI) in kg/m2. In pink are depicted the 90% prediction intervals (PIs) with conditional median from quantile regression forest (QRF); From left to right, the two visits for VHU and the baseline visit for UKB.
